# CAR T cell therapy in B-cell acute lymphoblastic leukaemia: Insights from mathematical models

**DOI:** 10.1101/2020.03.18.20038257

**Authors:** Odelaisy León-Triana, Soukaina Sabir, Gabriel F. Calvo, Juan Belmonte-Beitia, Salvador Chulián, Álvaro Martínez-Rubio, María Rosa, Antonio Pérez-Martínez, Manuel Ramirez-Orellana, Víctor M. Pérez-García

**Affiliations:** Department of Mathematics, Mathematical Oncology Laboratory (MOLAB), Universidad de Castilla-La Mancha, Ciudad Real, Spain; Faculty of Sciences, University Mohammed V, Rabat, Morocco; Department of Mathematics, Universidad de Cádiz, Puerto Real, Cádiz, Spain; Translational Research Unit in Paediatric Haemato-Oncology, Hematopoietic Stem Cell Transplantation and Cell Therapy, Hospital Universitario La Paz, Madrid, Spain; Paediatric Haemato-Oncology Department, Hospital Universitario La Paz, Madrid, Spain; Department of Pediatric Hematology and Oncology, Hospital Infantil Universitario Niño Jesús, Universidad Autónoma de Madrid, Madrid, Spain

**Keywords:** Mathematical modelling, Cancer dynamics, Immunotherapy, Tumor-immune system interactions, Mathematical oncology

## Abstract

Immunotherapies use components of the patient immune system to selectively target cancer cells. The use of CAR T cells to treat B-cell malignancies – leukaemias and lymphomas– is one of the most successful examples, with many patients experiencing long-lasting complete responses to this therapy. This treatment works by extracting the patient’s T cells and adding them the CAR group, which enables them to recognize and target cells carrying the antigen CD19^+^, that is expressed in these haematological tumors.

Here we put forward a mathematical model describing the time response of leukaemias to the injection of CAR T-cells. The model accounts for mature and progenitor B-cells, tumor cells, CAR T cells and side effects by incorporating the main biological processes involved. The model explains the early post-injection dynamics of the different compartments and the fact that the number of CAR T cells injected does not critically affect the treatment outcome. An explicit formula is found that provides the maximum CAR T cell expansion in-vivo and the severity of side effects. Our mathematical model captures other known features of the response to this immunotherapy. It also predicts that CD19^+^ tumor relapses could be the result of the competition between tumor and CAR T cells analogous to predator-prey dynamics. We discuss this fact on the light of available evidences and the possibility of controlling relapses by early re-challenging of the tumor with stored CAR T cells.

## 1. Introduction

Cancer immunotherapy approaches use components of a patient’s own immune system to selectively target cancer cells. Immunotherapies are already an effective treatment option for several cancers due to its selectivity, long-lasting effects, benefits on overall survival and tolerance [1].

Chimeric antigen receptor (CAR) T-cell therapy represents a major step in personalized cancer treatment, being the most successful type of immunotherapy. This therapy is predicated on gene transfer technology to instruct T lymphocytes to recognize and kill tumor cells. CARs are synthetic receptors that mediate antigen recognition, T cell activation, and costimulation to augment T cell functionality and persistence. For its clinical application, patient’s T cells are obtained, genetically engineered ex-vivo to express the synthetic receptor, expanded and infused back into the patient [2].

Clinical trials have shown promising results in end-stage patients with B-cell malignancies due to their expression of the CD19 protein [3]. CAR T cells engineered to recognize this antigen have lead to an early clinical response of up to 92% in Acute Lymphoblastic Leukemia (ALL) patients [4, 5, 6, 7]. Good results have been reported for large B-cell lymphomas [8, 9] and multiple myelomas [10]. These successes have led to the approval of CAR T therapies directed against CD19 for treatment of B-ALL and diffuse large B-cell lymphomas [3].

CAR T-related toxicities are the cytokine release syndrome (CRS), due to the release of cytokines during CAR T cell action, and immune effector cell associated neurotoxicity syndrome (ICANS). CRS symptoms including hypotension, pulmonary edema, multiorgan failure, and even death, are now better controlled using tocilizumab [11].

A variable fraction between 30% and 60% of patients relapse after treatment, probably due to persistence of CAR T-cells and escape or downregulation of the CD19 antigen [12]. The immunophenotype of post-CAR blasts in CD19^+^ relapses, is the same as preinfusion disease in most of the relapses, consistent with return of the pre-CAR immunophenotypic clone. Pre-CAR and post-CAR blasts typically show the same CD19 expression levels in flow cytometry. CAR T cells are able to kill blasts with low levels of CD19 expression. In fact, any CD19 level detectable by flow cytometry would trigger the action of CAR T cells. In contrast to CD19^+^ recurrences, CD19^−^ recurrences occur despite functional persistence and ongoing B-cell aplasia [12, 13].

There have been a large number of previous works devoted to the mathematical modelling of tumor-immune cell interactions, see for instance [14, 15, 16, 17, 18, 19] and references therein. Mathematical models have the potential to provide a mechanistic understanding of oncologic treatments and may help in finding optimized strategies improving treatment outcome [20, 21]. This is why CAR T cell treatments have attracted the interest of mathematicians in the context of gliomas [22, 23], melanomas [24] and B-cell malignancies [25, 26, 27].

In this work, we will describe mathematically the longitudinal dynamics of B cells, leukaemic clones and CAR T cells. The mathematical models will be shown to provide both a mechanistic explanation to the results of different clinical trials and formulas quantifying some of the observed phenomena. We will discuss some implications for CD19^+^ relapses and how it could be possible to control them by early rechallenging the tumor with CAR T cells.

## 2. Mathematical models and parameter estimation

### 2.1. Basic mathematical model

Our mathematical model accounts for the temporal evolution of several interacting cellular populations distributed into five compartments. Let *C*(*t*), *T* (*t*), *B*(*t*), *P* (*t*), and *I*(*t*) denote the nonnegative time-varying functions representing the number of CAR T cells, tumor (leukemic) cells, mature healthy B cells, CD19^−^ hematopoietic stem cells (HSCs), and CD19^+^ B cell progenitors (i.e. Pre-B, Pro-B and immature bone marrow B cells), respectively.

Our starting autonomous system of differential equations reads as

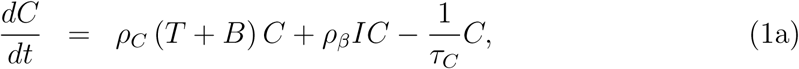

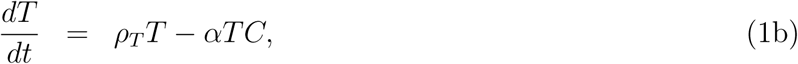

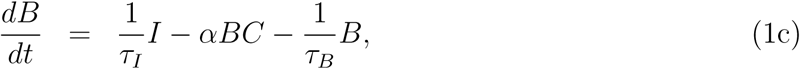

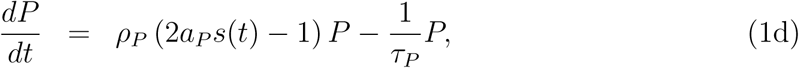

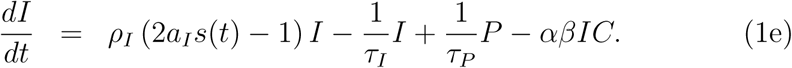

Equation (1a) involves two proliferation terms of CAR T cells due to stimulation by encounters with their target cells: either *T* (*t*), *B*(*t*) or *I*(*t*). The parameter *ρ*_*C*_ *>* 0 measures the mitosis stimulation after encounters with CD19^+^ cells that are disseminated throughout the entire body (mostly in the circulatory system). The parameter *ρ*_*β*_ = *βρ*_*C*_, where 0 *< β <* 1, accounts for the fact that immature B cells are located in the bone marrow and encounters with CAR T cells will be less frequent. The last term describes the decay of CAR T cells with a mean lifetime *τ*_*C*_.

In contrast with previous modelling approaches [25, 26, 27], we exclude a death term in the CAR T cell compartment due to interaction with target cells. The reason is that CAR T cells do not undergo apoptosis after killing the target cell [28, 29]. Also, different from those models, a *standard* proliferation term proportional to the population of CAR T cells is absent since these cells do not divide spontaneously [30]; instead their clonal expansion is directly dependent on the stimulation with the CD19 antigen.

Tumor cells [see Eq. (1b)] have a net proliferation rate *ρ*_*T*_ *>* 0 and die due to encounters with CAR T cells. The parameter *α* measures the encounter probability (per unit time and cell) of CAR T with CD19^+^ cells. Both *α* and *ρ*_*C*_ may, in general, be different from each other due to possible asymmetric cell interactions. Namely, if *α > ρ*_*C*_, this would imply that CAR T cells kill CD19^+^ target cells relatively faster than their own proliferation rate per target cell encountered. In contrast, if *α < ρ*_*C*_ then, on average, the killing process would be slower than the proliferation rate per target cell encountered by CAR T cells. For completeness, we will consider both cases via the unitless parameter *k* = *ρ*_*C*_*/α*. Other processes, encompassed by the term −*αTC*, would include target cell recognition, killing and detachment, which are relatively much faster than the complete *rendezvous* kinetics [31]. Our model also implicitly considers that both CD4^+^ T helper cells and CD8^+^ T cells have a similar cell killing capacity, although a more complex framework should regard the existing differences in the killing capacity of these two T lymphocytes [31, 32].

Equations (1c)-(1e), that involve B cells, consist of a compartment for CD19^−^ HSCs (i.e. *P* (*t*)) having an asymmetric division rate *a*_*P*_ and a differentiation rate 1*/τ*_*P*_ into a new compartment accounting for all of the other CD19^+^ differentiated states of bone marrow progenitor B cells (Pro-B, Pre-B, and immature cells) (embodied in *I*(*t*)). These cells constitute the source of mature B cells. Since all cells in the *I*(*t*) compartment already express the CD19^+^ antigen, they will be targets for the fraction of CAR T cells in the bone marrow, namely *βC*(*t*). Finally, mature B cells *B*(*t*), which cannot subsequently proliferate, are the terminal differentiation stage of these cells. They have a mean lifetime *τ*_*B*_, which is present in the last term of Eq. (1c). The structure of the two hematopoietic compartments is similar to that proposed in previous works on hematopoiesis models [33]. In line with those models, the signalling function *s*(*t*) can be assumed to be of the saturable form *s*(*t*) = 1*/* [1 + *k*_*s*_ (*P* + *I*)], with *k*_*s*_ *>* 0.

To describe CRS, let us define a variable *Y* (*t*) related to the cytokines released upon stimulation of CAR T cells by the antigens with a rate *ρ*_*Y*_ and cleared up with a rate 1*/τ*_*Y*_. ICANS will be related to the number of CAR T-cells infiltrating the central nervous system (CNS), and is expected to be proportional to the total number of CAR T-cells, with a proportionality constant *ρ*_*N*_, and removed at a rate 1*/τ*_*N*_. Thus, toxicity can be accounted for by means of the following equations

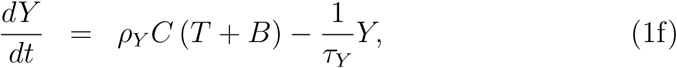

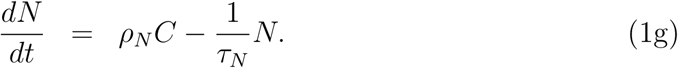

Notice that Eqs. (1a)-(1e) are uncoupled from Eqs. (1f)-(1g). A schematic summary of the biological processes encompassed by our basic mathematical model (1a-1g) is shown in Fig. 1.

**Figure 1:**
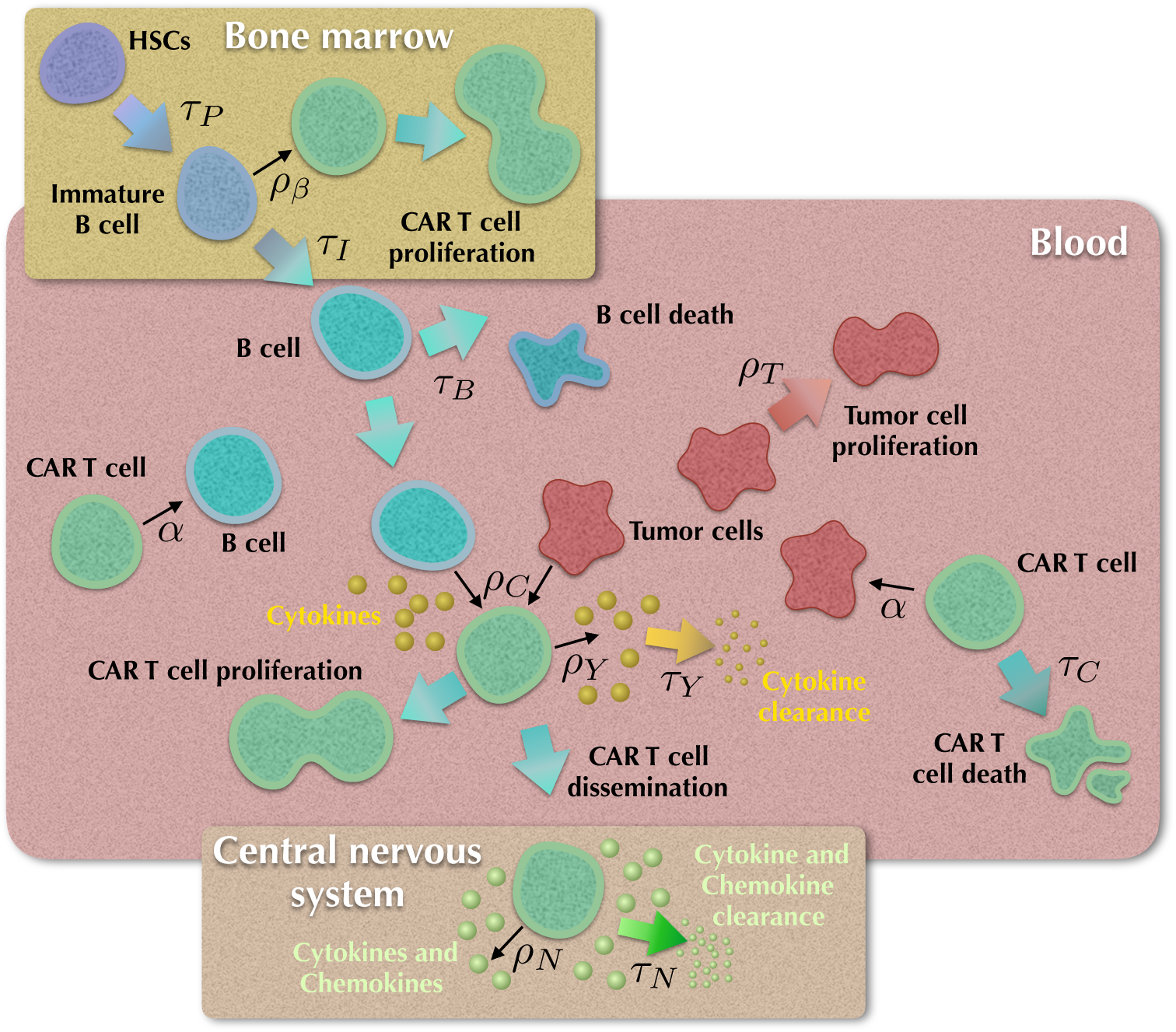
Processes included in the mathematical model (1). Mature B lymphocytes are generated from the CD19^−^ hematopoietic stem cells (HSCs) and through differentiation of immature CD19^+^ progenitors with characteristic lifetimes *τ*_*P*_ and *τ*_*I*_, respectively. CAR T cells are stimulated when meeting CD19^+^ B cells (normal, leukaemic or immature) with stimulation constants *ρ*_*C*_ and *ρ*_*β*_, and undergo apoptosis with a lifetime *τ*_*C*_. Tumor (leukaemic) cells proliferate with a rate *ρ*_*T*_. Both mature B and tumor cells are destroyed via encounters with the CAR T cells with a killing efficiency *α*. Cytokines are released with a rate *ρ*_*Y*_, that may result in acute toxicities (cytokine release syndrome) and are cleared at a rate 1*/τ*_*Y*_. A fraction of the CAR T cells disseminates and infiltrates the central nervous system. Typical neurological toxicities induced by CAR T cells are immune effector cell-associated neurotoxicity syndrome.

Appendix A shows some mathematical results on existence, uniqueness and positiveness of the solutions of system (1a)-(1e).

### 2.2. Reduced mathematical models

Equations (1a)-(1e) exclude different biological facts such as heterogeneity in the CAR T-lymphocyte subpopulations, the differential expression of the CD19 antigen across tumor and healthy B cells subclones, the role of regulatory T-cells, etc. However, it still has a large number of parameters to be determined. The contribution of the bone marrow Eqs. (1d)-(1e) is to account for the generation of new B-cells. Hence, to capture their role while simplifying the full system, we can compute the equilibria for Eqs. (1d)-(1e)

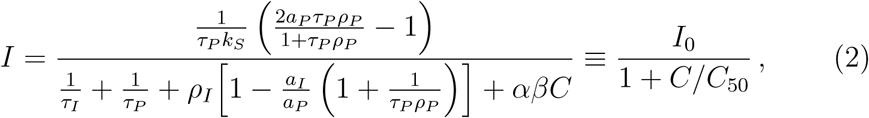

and assume (2) to hold for all times. This provides a suitable representation of the contribution of immature B cells in the bone marrow to the global disease dynamics. Then, Eqs. (1a)-(1e) reduce to the set

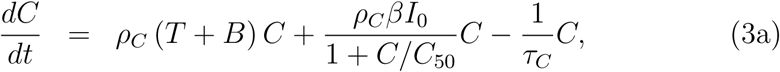

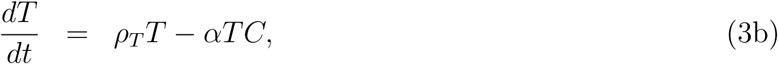

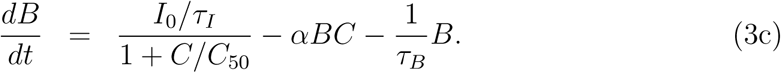

In the first weeks, after CAR T injection, the main contribution to the dynamics is the expansion of these cells and their effect on the healthy B and tumor cells. Thus, we may neglect the contribution of the hematopoietic compartments in Eqs. (3a) and (3c) to get

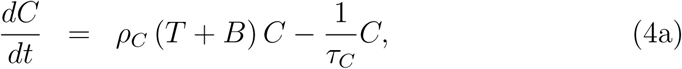

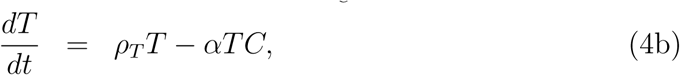

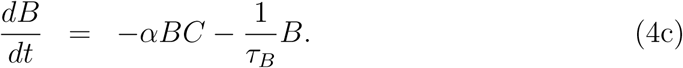

The study of existence and uniqueness of solutions, together with the stability of the critical points for both systems, are presented in Appendix B and Appendix C.

### 2.3. Parameter estimation

B-cell lymphocyte lifetime *τ*_*B*_ is known to be about 5-6 weeks [44]. These cells account for a variable fraction between 5% and 20% [45] of the total lymphocyte number [46] leading to *>* 10^11^ B-lymphocytes in humans.

ALLs are fast-growing tumors with proliferation rates *ρ*_*T*_ of the order of several weeks [33, 47]. Na ï ve CD8^+^ T cells are quiescent, their mean lifetime ranges from months to years, and enter the cell cycle following interaction with their antigen [48, 49]. These activated CD8^+^ T cells induce cytolysis of the target cells and secrete cytokines such as TNF-*α* and IFN*γ*. Following activation, most effector cells undergo apoptosis after two weeks, with a small proportion of cells surviving to become CD8^+^ memory T cells capable of longer survivals [48]. Recent works have reported longer survival values of about one month [13]. Thus, we will take the mean lifetime *τ*_*C*_ of CAR T cells in the range of 2-4 weeks.

To estimate the interaction parameter *α* we will use the fact that when measured by flow cytometry or qPCR, CAR T cells in children treated for ALL reached a maximum *in vivo* expansion around 14 days [35], that is a typical value observed in other clinical studies. Finally, the mitotic rate *ρ*_*C*_, related to the stimulatory effect of each encounter of T cells with the CD19^+^ cells, will be taken to be proportional to *α*. Parameter values employed in this paper are summarized in Table 1.

**Table 1:** Parameter values of relevance for model Eqs. (4)

| Parameter | Meaning | Value | Units | Source |
| --- | --- | --- | --- | --- |
| $\tau_B$ | B-lymphocyte lifetime | 30 – 60 | day | [44] |
| $\rho_T$ | Tumor growth rates | 1/30 – 1/60 | day<sup>-1</sup> | |
| $\tau_C$ | Activated CAR T cell lifetime | 14 – 30 | day | [48, 13] |
| $\rho_C$ | Mitotic stimulation of CAR T cells by $CD19^+$ cells | $(0.05 - 2) \times \alpha$ | day<sup>-1</sup> × cell<sup>-1</sup> | |
| $\alpha$ | Killing efficiency of CAR T cells | $\sim 10^{-11}$ | day<sup>-1</sup> × cell<sup>-1</sup> | Estimated from [35] |

## 3. Results

In this section, we present the results obtained from systems (3) and (4). We start by analyzing (4):

### 3.1. Mathematical model (4) describes post CAR T cell injection dynamics

Firstly, we numerically studied the dynamics of the system post-CAR T cell injection as described by Eqs. (4). Figure 2 shows a typical example. During the first two months simulated, CAR T cells expanded evidencing a peak at about two weeks post-injection, before their numbers stabilized and start decreasing. Both the tumor and B-cell compartments experienced a continuous decrease towards undetectable values representing the dynamics of a patient without residual disease. The expansion of the CAR T population was exponential, increasing by several orders of magnitude [see Figure 2(b)], in line with reported clinical experience and patient datasets [13].

**Figure 2:**
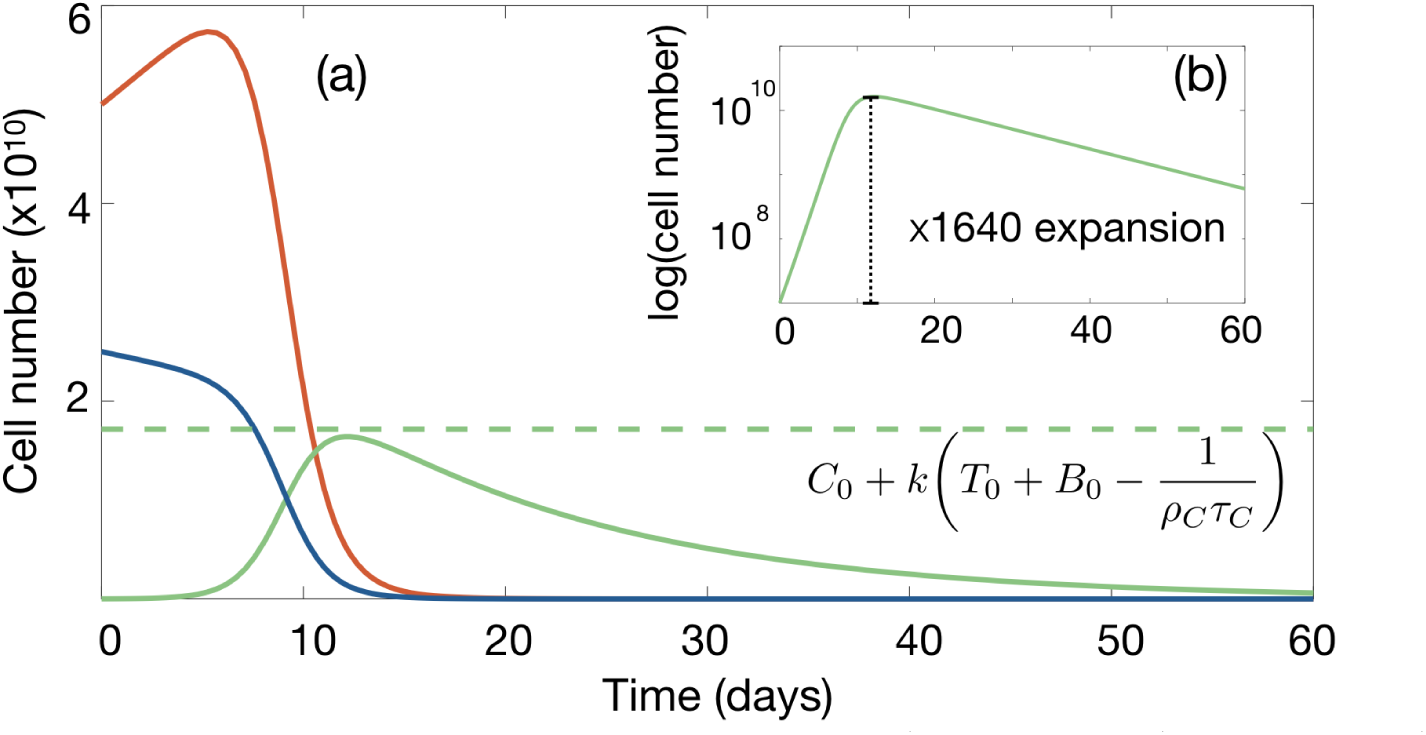
Typical dynamics of tumor (red curve), B-cell (blue curve) and CAR T cell (green curve) compartments according to Eqs. (4). (a) Simulations for parameters *α* = 4.5×10^−11^ day^−1^cell^−1^, *τ*_*C*_ = 14 days, *ρ*_*T*_ = 1/30 day^−1^, *ρ*_*C*_ = 0.25*α, τ*_*B*_ = 60 days and injected cells *C*_0_ = 10^7^ corresponding, to 5×10^5^ cells per kg for a 20 kg child. Also, *T*_0_ = 5×10^10^ and *B*_0_ = 2.5×10^10^, which correspond to typical values after lymphodepleting chemotherapy. (b) Logarithmic plot of the CAR T cell population.

### 3.2. The number of injected CAR T cells does not affect treatment outcome, but the stimulation rate does

We next studied the dynamics of Eqs. (4) under different numbers of injected CAR T cells. A typical example is displayed in Figure 3(a). The change of one order of magnitude in the initial CAR T cell load resulted in minor changes in the maximum expansion achieved (of around 6%). A reduction in the time to peak expansion *in-silico* of about 3 days was observed. However, the persistence of CAR T cells was not affected by their initial load.

**Figure 3:**
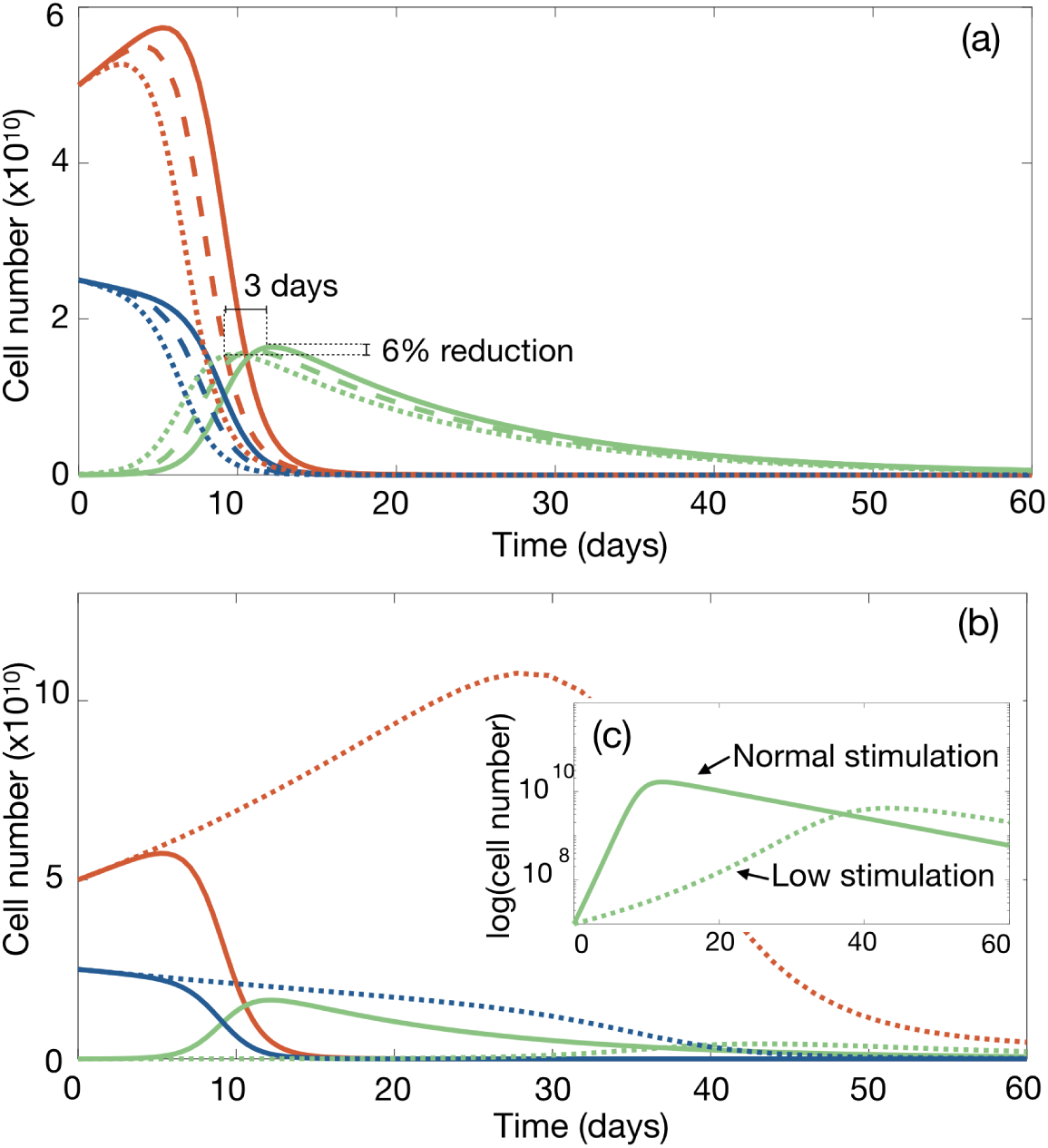
The number of injected CAR T cells does not affect treatment outcome, but the stimulation rate does. (a) Dynamics of tumor (red line), B-cells (blue line) and CAR T cells (green line) according to Eqs. (4) for the virtual patient of Fig. 2 subject to injections of 5 × 10^5^ cells/kg (solid lines), 15 × 10^5^ cells/kg (dashed lines) and 45 × 10^5^ cells/kg (dotted lines). (b,c) Dynamics for stimulation rates *ρ*_*C*_ = 0.25*α* (solid line) and *ρ*_*C*_ = 0.05*α* (dotted lines). (c) CAR T cells expansion in log scale.

It has been described that when the number of CAR T cells seeded is small, the therapy can fail [34]. However, the injection of low CAR T cell numbers is typically done when these cells do not expand well *in-vitro*, that could be the result of a low stimulation capability. We simulated *in-silico*, the effect of a reduction in the growth efficiency of the cells (the stimulation rate *ρ*_*C*_) and the dynamics was substantially affected [see Fig. 3]. A reduction of the stimulation efficiency in the CAR T cells led to a slower growth of this population *in-silico*, resulting in a tumor growth reaching unacceptable levels for almost two months without any clinical response.

### 3.3. Maximum expansion of CAR T cells in-vivo and CRS

System (4) is amenable for finding useful analytical and semi-analytical expressions, which are all derived in Appendix D.

At time *t*_max_, which typically occurs within 2-4 weeks after injection of the CAR T cells, a first maximum in their number, denoted by *C*_max_ ≡ *C*(*t*_max_), is achieved during the expansion phase. The value of *t*_max_ can be calculated from the implicit relation

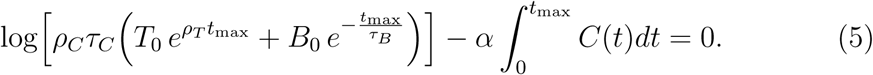

Furthermore, it is possible to estimate the maximum number of CAR T cells *C*_max_, which is approximately given by

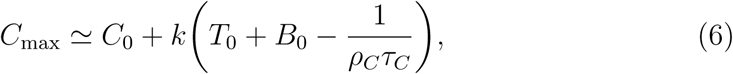

where *C*_0_, *T*_0_ and *B*_0_ are the initial conditions for the CAR T, tumor and B cells (they are assumed to be positive constants), respectively. Thus, the maximum number of CAR T cells that can be achieved during the first expansion phase will be related to the initial populations *T*_0_ and *B*_0_ multiplied by the ratio *k*. Notice also that, in practice, the contribution of *C*_0_ is much smaller than the second term in Eq. (6) and can be ignored, evidencing that the initial number of injected CAR T cells does not affect the peak, although it does contribute in (5) when computing *t*_max_. Both results (5) and (6) provide an analytical justification of the numerical results discussed for Eqs. (4) in Section 3.2.

It is easy to obtain explicit formulas to compute the tumor and B cell loads in patients at time *t*_max_. They are given by

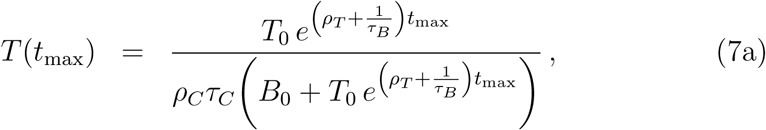

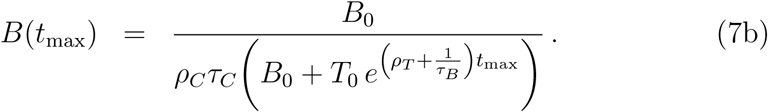

Alternatively, if *T* (*t*_max_) and *B*(*t*_max_) are available, together with constants *T*_0_, *B*_0_, *τ*_*C*_ and *τ*_*B*_, then parameters *ρ*_*C*_ and *ρ*_*T*_ can be estimated, which is currently both important and challenging from a clinical viewpoint.

Since toxicity, accounted for by Eqs. (1f) and (1g), depends on the maximum CAR T cell number one would expect a smaller ratio *k* to lead to lower toxicities. Figure 4 shows the linear dependence of the maximum number of CAR T cells on *ρ*_*C*_ as obtained from simulations of Eqs. (4), that is well approximated by Eq. (6). The above result points out to a proportionality dependence between the total tumor load and the severity of the CRS syndrome. In fact, a strong correlation between the severity of CRS and disease burden at the time of CAR-T cell infusion has been noted in multiple clinical trials of CAR-T cell therapy of hematological malignancies [35, 36, 37, 38].

**Figure 4:**
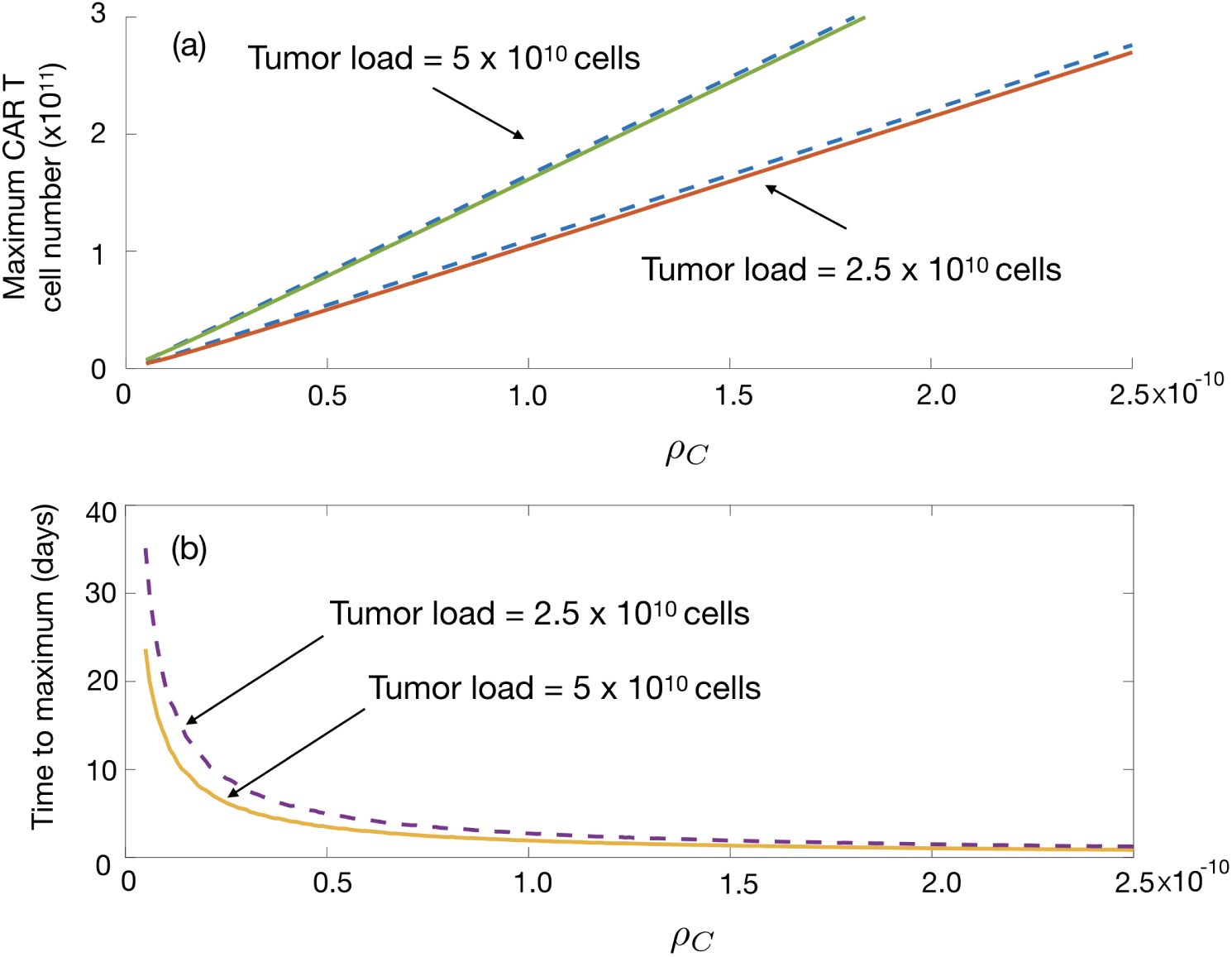
Dependence on *ρ*_*C*_ of the maximum number of CAR T cells and the time *t*_max_ taken to achieve the maximum. Common parameters for all plots are as in Figure 2: *α* = 4.5×10^−11^ cell^−1^ day^−1^, *τ*_*C*_= 14 days, *ρ*_*T*_ = 1/30 day^−1^, *τ*_*B*_ = 60 days and initial cell numbers *C*_0_ = 10^7^ cells, *B*_0_= 2.5 × 10^10^ cells. (a) Maximum value number of CAR T cells obtained for initial tumor loads of 5 × 10^10^ cells (red) and 2.5 × 10^10^ cells as a function of *ρ*_*C*_. Solid line indicates the results obtained from Eqs. 4 and the dashed line the upper bound given by Eq. (6). (b) Time to maximum value of CAR T cells for different tumor loads computed from Eq. (5).

### 3.4. CAR T cell persistence depends on the T cell mean lifetime

The recent clinical study [13] showed much longer persistence of the CAR T cells when their mean lifetime was increased to *τ*_*C*_ = 30 days, larger than the more common value *τ*_*C*_ = 14 days. We simulated the dynamics of Eqs. (4) for both values of *τ*_*C*_. An example is shown in Fig. 5. While the B-cells and tumor cells exhibited a similar behavior, CAR T cells showed a much longer persistence in line with the clinical observations.

**Figure 5:**
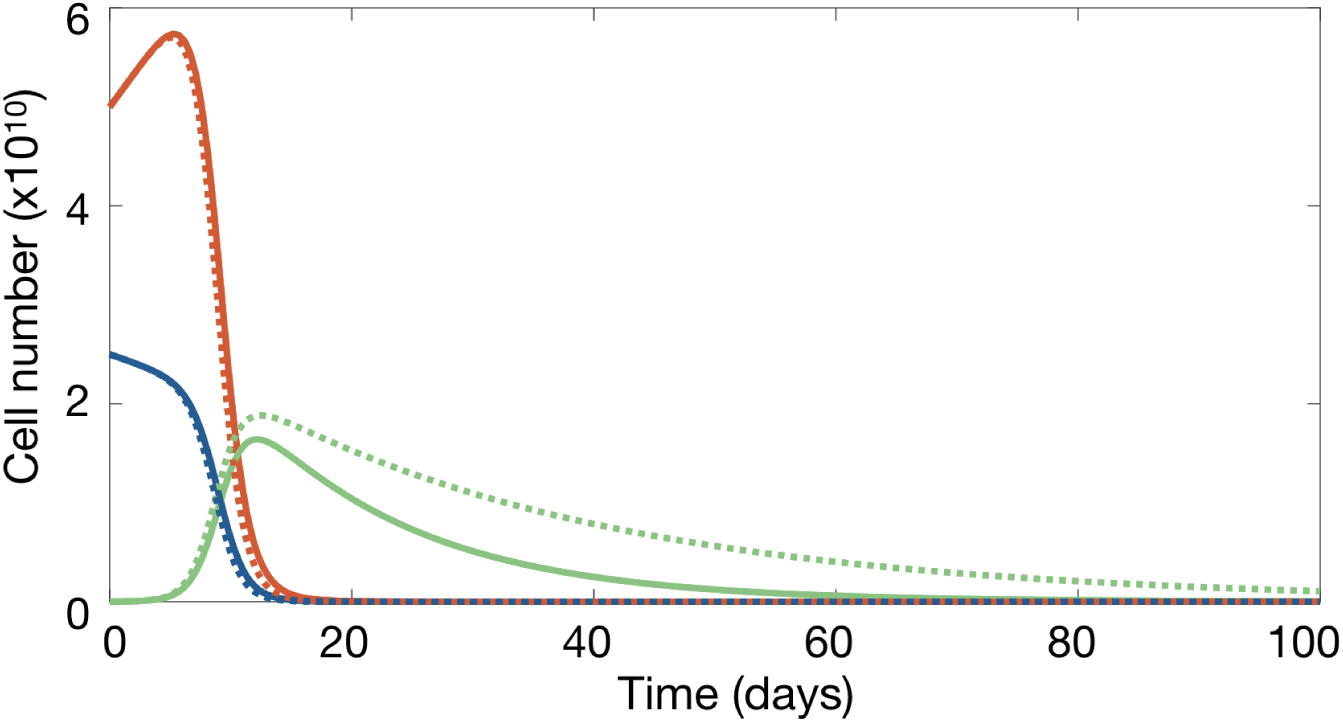
CAR T cell persistence when varying its lifetime *τ*_*C*_. Dynamics of the tumor (red), B-cell (blue) and CAR T cell (green) according to model Eqs.(4). Solid and dashed curves correspond to *τ*_*C*_ = 14 days and *τ*_*C*_ = 30 days, respectively, with the rest of parameters and initial data as in Figure 2.

#### 3.5. CD19^+^ relapses could be a dynamical phenomenon

We performed simulations of Eq. (4) for longer timescales (with parameters as in Fig. 2) and observed a long-time relapse (see Fig. 6) at about one year after infusion, in what would be a CD19^+^ relapse. Tumor growth continued for several months but finally there was an outgrowth of CAR T cells after the relapse that was able to control the disease. This is an important nonlinear dynamical phenomenon that could help explain some CD19^+^ relapses.

**Figure 6:**
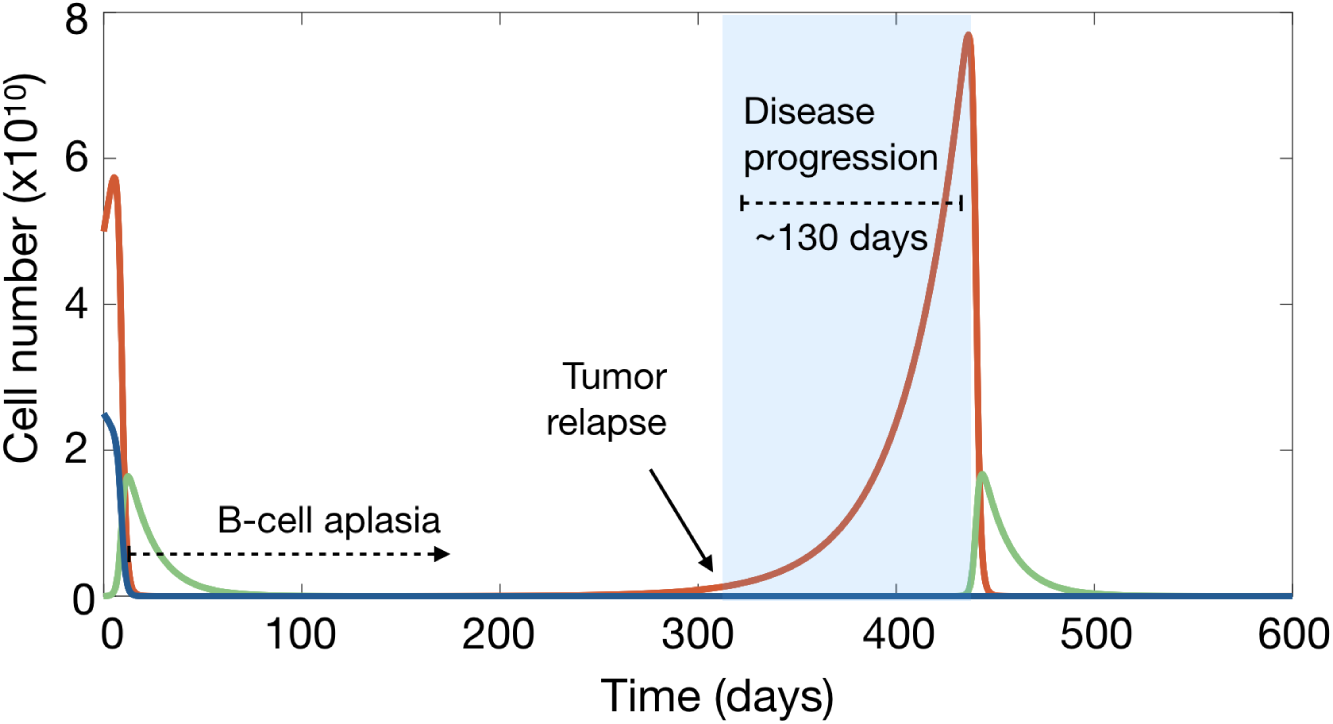
CD19^+^ relapses could be a dynamical phenomenon. Longtime dynamics of Eqs. (4) for tumor (red), B (blue) and CAR T (green) cells in the time interval [0,600] days, displaying a CD19^+^ relapse as the result of predator-prey type dynamics *in-silico*. Parameters are as in Figure 2. The shaded area indicates the time interval in which the disease would be progressing without further interventions. Notice the subsequent emergence of CAR T cells after the CD19^+^ cell relapse.

When *B*∼0, as it happens after CAR T cell expansion, Eqs. (4) become the well-known Lotka-Volterra predator-prey mathematical model. That model gives rise to periodic oscillations corresponding to ecological cycles that have been observed both in ecosystems [39] and in experimental models [40]. In our present case, the period of the cycles would be related to the tumor relapse time. This period has a complex dependence on a conserved quantity, the energy *E* [41], given by

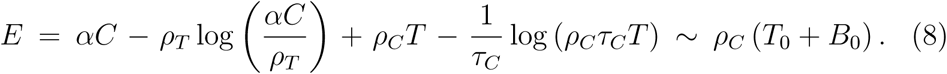

For values of the energy *E* ≫1, there is an asymptotic formula [42], that can be applied in our case yielding the period 𝒯 of oscillations

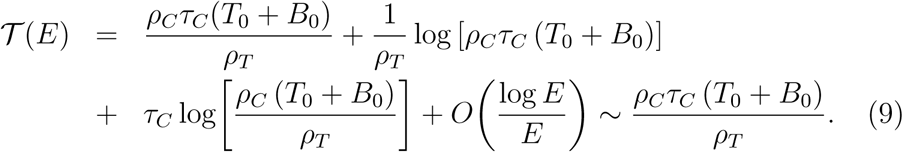

Because of the approximations involved, Eq.(9) should only be taken as an order-of-magnitude estimate for the tumor relapse time. However, it is interesting that longer lifetimes of the CAR T cells resulted in longer relapse times according to Eq. (9). This could be the reason why so few CD19^+^ relapses were observed in the recent trial [13], with *τ*_*C*_ = 30 days, much longer than the more common value *τ*_*C*_ ∼ 14 days.

A very intriguing question is if those relapses could resolve spontaneously due to the predator-prey type competition between the CAR T and tumor cells. Owing to the long progression time observed this could pass unnoticed since after progression other therapeutical actions would be taken, such as hematopoietic transplants, before letting the CAR T to appear again.

Although our simulations point out to a potentially interesting scenario, the model given by Eqs. (4) is not a good one to study long-time phenomena. One of the missing biological processes in Eqs. (4) is the potential contribution of B-cell production in the bone marrow from CD19^−^ hematopoietic stem cells to the maintenance of a pool of CAR T cells. Thus, to get a more realistic insight on the dynamics, we simulated Eqs. (3) for the same virtual patients and educated guesses for the parameters *β* = 0.1, *τ*_*I*_ = 6 days and different values for the production of B cells in the bone marrow embodied by *I*_0_.

One set of examples is shown in Figure 7. The more realistic model given by Eqs. (3) still presents a first relapse at a time that is independent of the choice of the flux *I*_0_. However this parameter influenced the post-relapse dynamics. For values of *I*_0_ smaller than approximately 10^7^ cells/day, which is the typical number of injected CAR T cells [34], there were no substantial changes in the dynamics with subsequent relapses following a periodic pattern. Larger values of *I*_0_ led to relapses of B cells before the relapse of tumor cells and later of CAR T cells. Also, the relapse dynamics was in line with damped oscillations, with both tumor and CAR T cell relapses having smaller amplitudes. Interestingly, our model Eqs. (3) predict that a significant increase of B cells could be used as a potential clinical biomarker indicative of subsequent tumor relapse.

**Figure 7:**
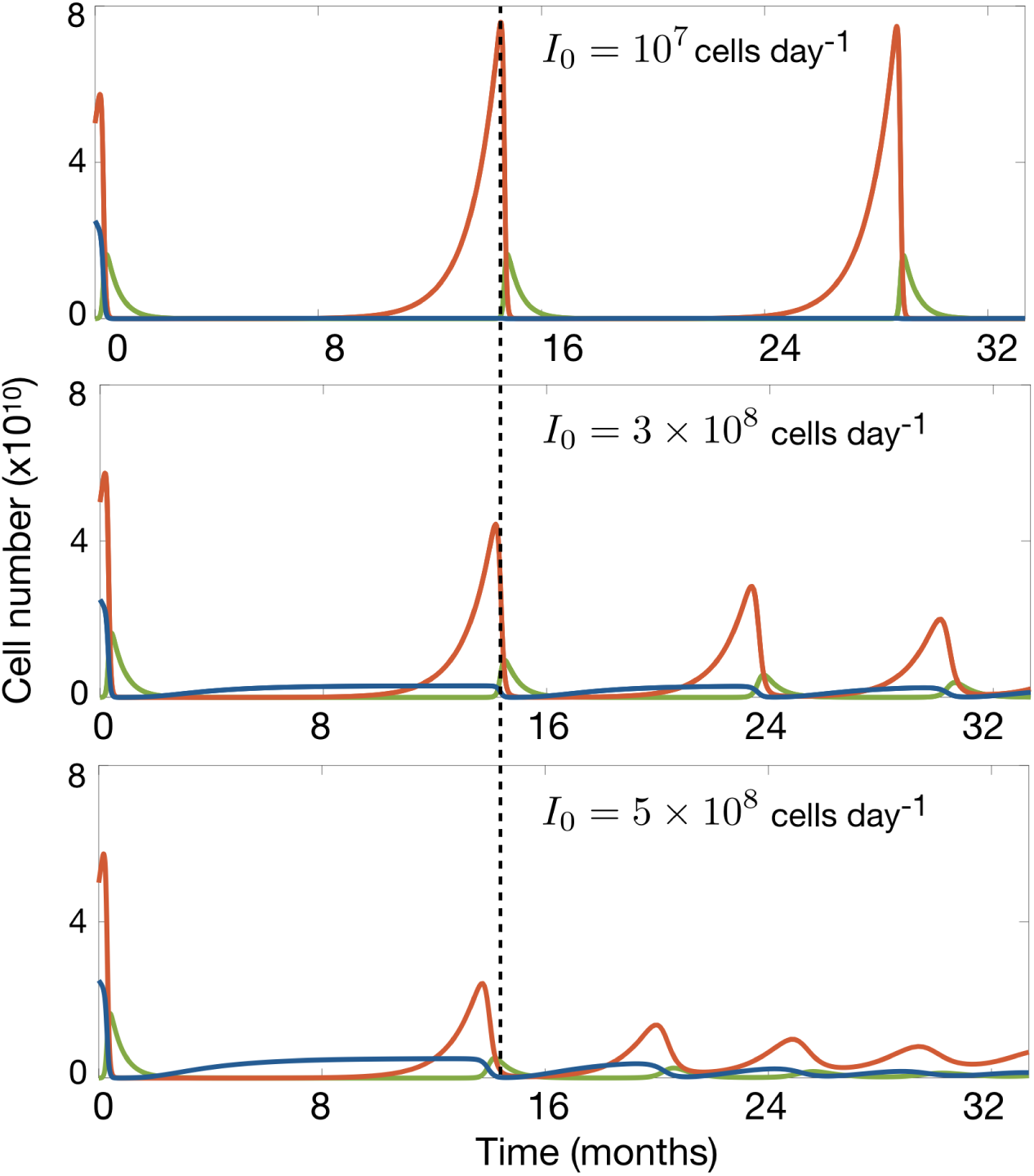
Long-time dynamics of virtual patients ruled by Eqs. (3). Parameters are *α* = 4.5×10^−11^ cell^−1^ day^−1^, *β* = 0.1, *τ*_*C*_ = 14 days, *τ*_*B*_ = 60 days, *ρ*_*T*_ = 1*/*30 day^−1^, *ρ*_*C*_ = 0.25*α, C*_50_ = 10^9^ cells, *τ*_*I*_ = 6 days. The subplots show the dynamics of the tumor (red), B-cell (blue) and CAR T cell (green) compartments. (a) Case *I*_0_ = 10^7^ cell day^−1^. (b) Case *I*_0_ = 3×10^8^ cell day^−1^, (c) Case *I*_0_ = 5 × 10^8^ cell day^−1^.

### 3.6. CART cell reinjection may allow to control relapse severity

In the framework of our modelling approach tumor relapses would be transient. However, the potential long duration of those relapses would require further intervention to avoid potential harm to the patient due to the tumor effect. An interesting question is if one could control relapses by acting on the tumor by reinjecting CAR T cells and what would be the right timings and doses for that intervention.

Using the mathematical model (3), we simulated the reinjection of *C* = 10^7^ CAR T cells at different times: before relapse (*t* = 300 days), on relapse (*t* = 360 days), and after relapse (*t* = 415 days) and compared the outcome with the case without reinjection. An example is shown in Figure 8(a,b).

**Figure 8:**
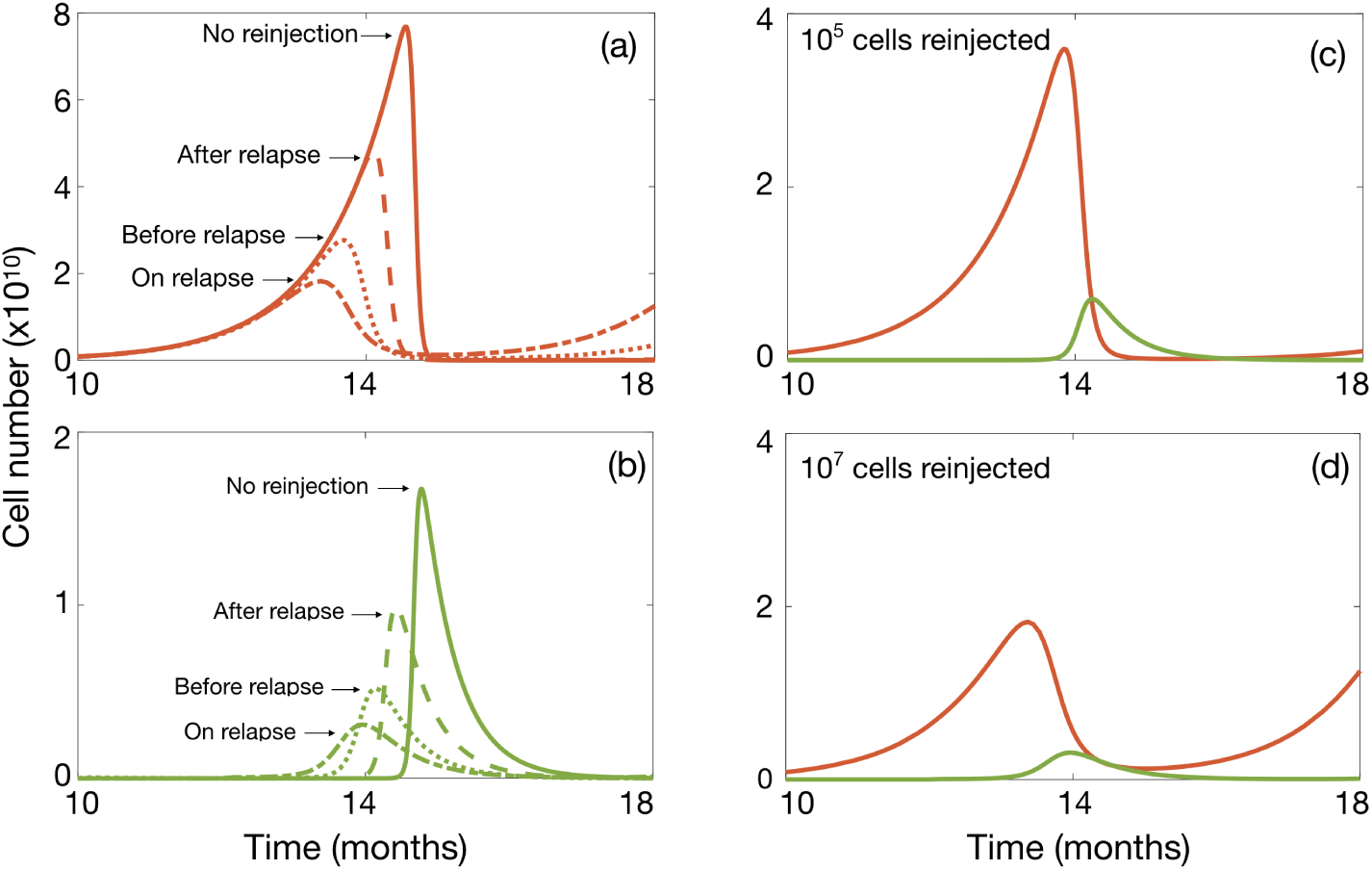
CAR T cell reinjection may allow to control relapse severity. Simulations of Eqs.(4) for parameter values *α* = 4.5×10^−11^ cell^−1^ day^−1^, *β* = 0.1, *τ*_*C*_ = 14 days, *τ*_*B*_ = 60 day, *ρ*_*T*_ = 1*/*30 day^−1^, *ρ*_*C*_ = 0.25*α, C*_50_ = 10^9^ cells, *τ*_*I*_ = 6 days, and *I*_0_ = 2×10^5^ cell day^−1^. All subplots show the dynamics of tumor (red) and CAR T (green) cells upon reinjection of CAR T cells. Cases (a) and (b) display the dynamics of (a) tumor and (b) CART cells, respectively, for doses of *C* = 10^7^ cells administered at times: *t* = 300 days (dash-dot line), *t* = 360 days (dotted line) and *t* = 415 days (dashed line) in comparison with the dynamics without reinjection (solid line). Subplots (c) and (d) illustrate the combined dynamics of tumor and CART cells after reinjection of: (c) *C* = 10^5^ cells and (d) *C* = 10^7^ cells at *t* = 360 days.

Significant reductions of both the peak tumor cell number and relapse duration were obtained, the best results being attained when reinjection was performed on relapse. Thus, our *in-silico* results suggest that the early reinjection of CAR T cells in a CD19^+^ B-leukemia relapse could reduce the disease burden and help in early control of the disease. This has interesting implications since, after relapse detection, CAR T preparation requires blood extraction, apheresis, T cell modification and expansion *ex-vivo*, and finally patient infusion. In the clinical practice this process takes between three to six weeks. From the practical point of view, a possibility to perform a faster action after tumor cell identification would be to freeze and keep part of the CAR T cells obtained initially so that they could be reinjected and aid in an early control of the disease.

We also studied the effect of the number of CAR T cells injected at the optimal time. An example is shown in Figure 8(c,d). The effects of a very small infusion of *C* = 10^5^ cells is compared with a more standard dose of *C* = 10^7^ cells. The number of T cells injected had an influence on the outcome. This was different from our previous observation that the number of CAR T cells injected initially did not affect the treatment outcome. The reason is that, initially, there are many targets, both tumor and B cells, providing support for a huge expansion of the CAR T population. However, on relapse, the target population is smaller and a larger initial number of CAR T cells helps in making the expansion process faster.

### 3.7. Model (3) predicts an scenario leading to a zero number of tumor cells

The analysis of the nonnegative equilibrium points of system (3), presented in Appendix C, shows the possibility to reach *T* = 0 starting from a nonzero tumor cell population. Using the parameters given in Table 1, we observe that there exist ranges for the ratio *k* and the bone marrow B cell production *I*_0_ where one of the equilibrium points, *P*_3_ (a focus), is asymptotically stable for different values of *α*. Hence, one of its associated eigenvalues will be real (see Figure C.12 in Appendix C) while the other two eigenvalues will be complex conjugate (see Figure C.13 in Appendix C). Figure 9 illustrates an example with different initial conditions and the same set of parameters for them. It should be pointed out that in all cases shown in Figure 9, both the CAR T and the B cells remain at nonzero levels (of the order of 5×10^8^ and 10^9^, respectively) when the tumor cell population effectively becomes extinct (*T*≪1.)

**Figure 9:**
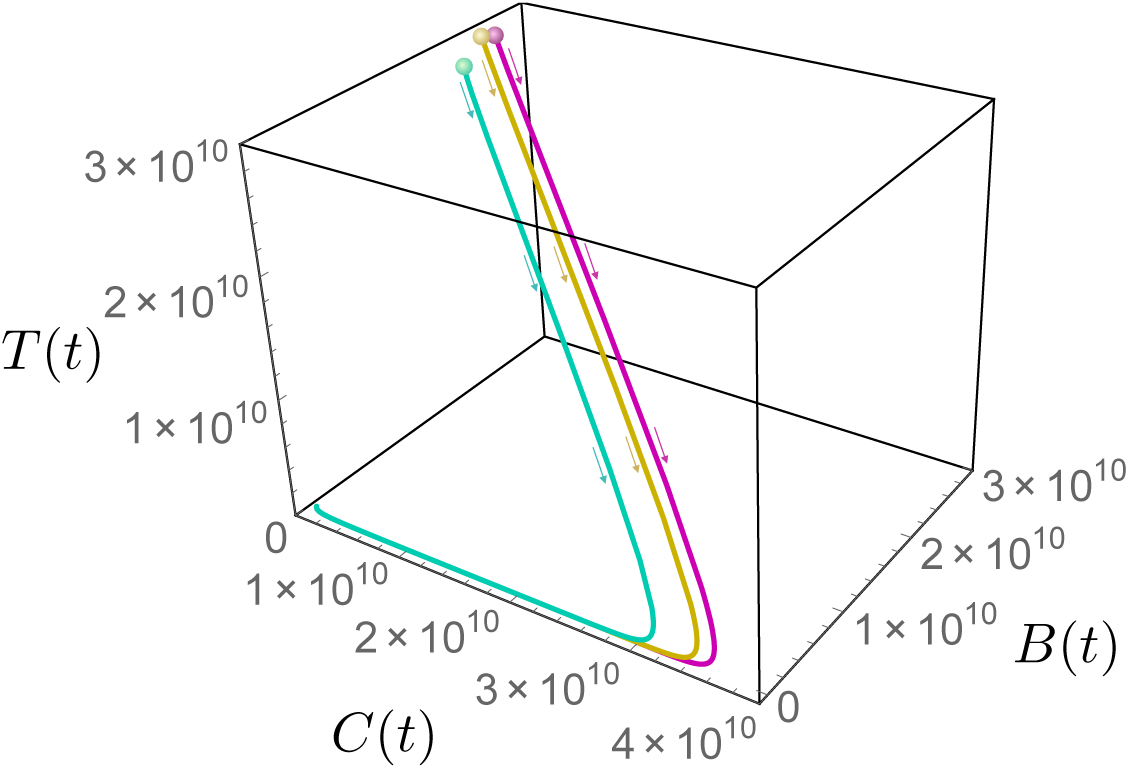
Routes to tumor cell extinction. Phase portrait of system (3) showing three orbits with different initial conditions (colored dots) that lead to tumor extinction. Parameters for all orbits are *α* = 5×10^−11^ cell^−1^ day^−1^, *β* = 0.1, *τ*_*C*_ = 20 days, *τ*_*B*_ = 40 day, *ρ*_*T*_ = 1*/*45 day^−1^, *ρ*_*C*_ = 0.7*α, C*_50_ = 10^9^ cells, *τ*_*I*_ = 2.4 days, and *I*_0_ = 2 × 10^8^ cell day^−1^.

These results are interesting as they suggest that there is a range of biologically relevant parameters where the tumor may eventually disappear. Our simulations indicate that the larger *I*_0_, *k* and *α*, the more likely it is to reach *T* = 0. Particularly, if *I*_0_ is increased, this would imply both higher production of B and CAR T cells, due to the contribution of immature B cells from the bone marrow, resulting in a larger probability to eradicate the tumor. This scenario provides another proof of concept that the mathematical model put forward here can be useful in the clinic and may trigger new exploratory pathways.

### 3.8. Sensitivity analysis

A sensitivity analysis was carried out to identify the model parameters displaying a greater influence on the equilibria for CAR T, tumor and B cells are *I*_0_, *α* and *k*. To do so, we calculated the first-order sensitivity coefficient using Sobol’s method [52] to measure the fractional contribution of a single parameter to the output variance. Using a priori information on the parameters, we defined the distribution functions in the table shown in Figure 10. We generated a set of parameters of size 1000 to calculate the sensitivity indices. The results of the sensitivity analysis of Eqs. (3) are depicted in Figure 10.

**Figure 10:**
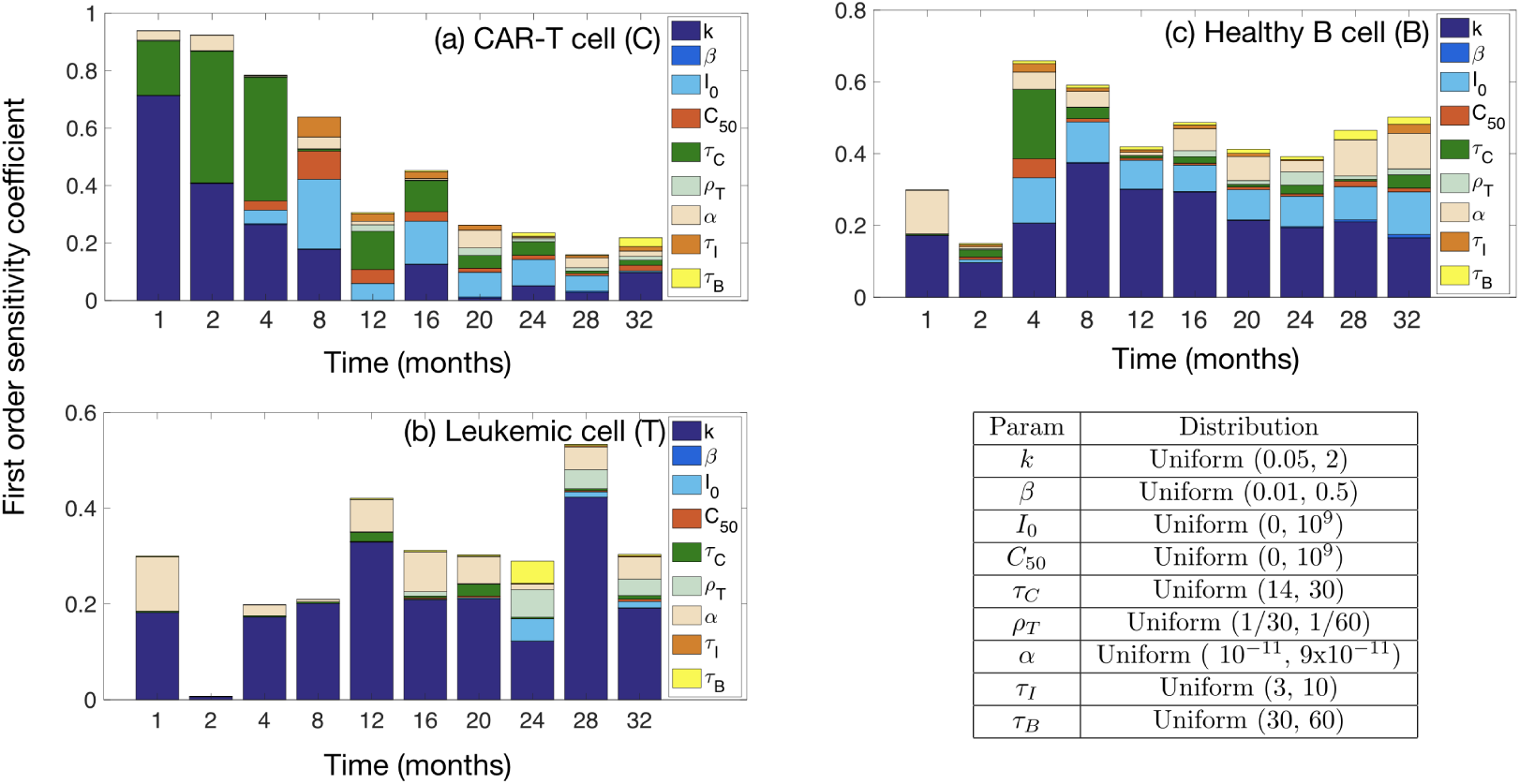
Sensitivity analysis of system (3) to identify the influence of the model parameters on the solutions for (a) CAR T, (b) tumor and (c) B cells. The parameter ranges studied and distributions used are displayed in the lower-right table.

The results show that the parameters having a larger influence on the solutions for CAR T, tumor and B cells are *k, α, τ*_*C*_ and *I*_0_. However, their impact varies depending on the specific cell compartment and time. For CAR T cells, *k* and *τ*_*C*_ are the most relevant during the first four months after injection. For longer times, *I*_0_ becomes the most important one. For tumor cells, *k* and *α* are the most influential parameters, both during the first weeks of the CAR T cell treatment and later during relapse. For healthy B cells, *k* and *α* are the most relevant parameters during the first weeks, but later on, on relapse, *I*_0_ also becomes important. Such parameter dependences are also suggestive in order to target specific mechanisms that would allow partial control over them.

## 4. Discussion

Cancer immunotherapy with CAR T cells is a promising therapeutic option already available for B cell haematological cancers. There has been a growing interest on the mathematical descriptions of immunotherapy treatments in cancer, particularly aimed at CAR T cells [25, 26, 27], but none of them have addressed the specificities of CAR T-based immunotherapies for the case of acute lymphoblastic leukaemias (ALL) on the light of available clinical experience.

In this work, we put forward a mathematical model incorporating the main cell populations involved in the growth of ALL. The model included not only leukaemic clones and CAR T cells, but also the hematopoietic compartment that would be responsible for the persistence of CAR T cells by the continuous generation of CD19^+^ progenitors from CD19^−^ stem cells.

One simplified version of the full model already allowed us to describe the clinical evolution of B ALL in the first months after CAR T injection yielding explicit formulas of clinical added value such as the maximum number of CAR T cells that can be reached. Also, it provided a rational support to several clinical observations. Interestingly, the model predicted the possibility of CD19^+^ relapses being dynamical phenomena resembling predator-prey oscillations. The more complex mathematical models were used to confirm this dynamic and to further give support for therapeutically rechallenging the tumor with CAR T cells in CD19^+^ relapses.

Although our model is shown to provide a somewhat complete description of the response to CAR T cell treatments, there are several limitations in extending our analysis to longer times and to the study of resistances. First of all, we did not study the case of CD19^−^ relapses. That study would require a different type of modelling in line with previous mathematical frameworks accounting for the development of other types of resistances under the evolutive pressure of treatments [43]. Also, our description of CD19^+^ relapses was based on a continuous model, that is not designed to faithfully capture the regime where the numbers of predators (CAR T cells) and preys (tumor and B cells) are low. It has been discussed that in those scenarios the situation is significantly more complex and may require different types of approaches, such as those based on stochastic birth-death processes or even fully discrete models accounting for different body compartments. However, the fact that the bone marrow compartment would provide a continuous flux of B cells leading to the maintenance of a pool of activated CAR T cells, may keep the system out of the very low densities that could make the continuous model fail.

B-cell lymphomas are usually treated with rituximab to provide a permanent lymphodepletion affecting also HSC. Although lymphomas are different diseases, it is interesting to mention that CAR T cells will be expected to persist for shorter times in that scenario due to the lack of the continuous stimulation for maintenance of a pool of those cells in the bone marrow. Thus, one anticipates that treatments would be less effective for those tumors.

The results obtained using the reduced mathematical model show that the number of CAR T cells initially injected does not affect the subsequent dynamics. Since at the outset CAR T cells do have a huge *in-vivo* target pool, including tumor and healthy B cells allowing them to expand, even small doses of properly functioning immune cells would lead to a response. Thus, according to our modelling approach, it may be better to store (freeze) part of the cells generated so that they could be ready for later tumor rechallenging in case of a CD19^+^ relapse. There, the combination of a fast action after the detection of the disease and the injection of a substantial number of CAR T cells would be clinically relevant according to our mathematical model-based predictions. The reason is that a prompt action would allow both for a reduced growth of the disease and for a smaller toxicity of the disease, thus reducing risks for the patient such as CRS and ICANS. The rationale behind the injection of larger CAR T loads on relapse is that the target population would be smaller in general than at the start of the treatment. Moreover, our model implies that a periodic treatment with CAR T cells to avoid relapse would be quite ineffective, since they would not be expected to expand well unless there is a substantial target population.

Our mathematical model allowed us to obtain an estimate for the very relevant parameter of the relapse time, that would be the optimal time to perform the re-injection of CAR T cells. The estimation obtained by Eq. (9) shows that the relapse time depends on the parameters related to CAR T cells (*ρ*_*C*_ and *τ*_*C*_), the growth rate *ρ*_*T*_ of tumor cells and their density at the beginning of treatment. In the framework of our continuous model all tumors experience a relapse, however longer relapse times may correspond in some simulation runs to tumors having intermediate densities unrealistically low for very long times. Thus, we may expect that tumors with quite long relapse times, according to our modelling approach, to never relapse.

We may act therapeutically on *ρ*_*C*_ and *τ*_*C*_ by designing CAR T cells with higher stimulation ratios and longer persistence. However, it is important to note that the first parameter would be expected to influence the initial treatment toxicity (CRS and ICANS), thus probably increasing the second would be a best option, perhaps by increasing the fraction of CD8^+^ memory T cells in the CAR T cell pool. Contradictory evidences have been found in this respect, although in this case the endpoint to consider would be the number of CD19^+^ tumor relapses.

Our mathematical model also allows us to pose another interesting hypothesis. B-ALL has been a field where substantial progresses have been achieved by designing initial intensive treatment regimes combining different types of cytotoxic chemotherapies. On the basis of our mathematical model, and leaving aside the important economical costs, one would expect that substantially less aggressive chemotherapy regimes could be quite effective after CAR T cell injection to eliminate the residual disease (mainly by greatly reducing the *ρ*_*C*_ parameter). That strategy would also be beneficial to control CD19^−^ relapses. In that case one should balance the side effects of current protocols versus a combination of CAR T cells with a reduced chemo infusion. The main limitation of this approach would be the sustained B-lymphodepletion provoked by the immunotherapy treatment, but one can envision autologous B stem cell transplants after *in-vitro* treatment with CAR T cells to select for CD19^−^ HSCs.

In conclusion we have put forward a mathematical model describing the response of acute lymphoblastic leukemias to the injection of CAR T cells. Our theoretical framework provided a mechanistic explanation of the observations reported in different clinical trials. Moreover, it also predicted that CD19^+^ tumor relapses could be the result of the competition between tumor and CAR T cells in an analogous fashion to predator-prey dynamics. As a result, the severity of relapses could be controlled by early rechallenging the tumor with previously stored CAR T cells.

## Data Availability

The manuscript has not data.

## Acknowledgement

This work has been partially supported by the Junta de Comunidades de Castilla-La Mancha (grant number SBPLY/17/180501/000154), the James S. Mc. Donnell Foundation (USA) 21st Century Science Initiative in Mathematical and Complex Systems Approaches for Brain Cancer (Collaborative award 220020450), Junta de Andaluca group FQM-201, Fundacin Espaola para la Ciencia y la Tecnologa (FECYT, project PR214 from the University of Cdiz) and the Asociacin Pablo Ugarte (APU). OLT is supported by a PhD Fellowship from the University of Castilla-La Mancha research plan.

## Appendix A. Basic properties of the complete mathematical model: system (1)

We state the following proposition:

### Proposition 1.

*For any nonnegative initial data* (*C*(0), *T* (0), *B*(0), *P* (0), *I*(0)) *and all parameters of the initial value problem given by Eqs. (1a)-(1e) being positive, the solutions for C*(*t*), *T* (*t*), *B*(*t*), *P* (*t*), *and I*(*t*) *exist for all t >* 0, *are unique and nonnegative*.

**Proof**. We first show nonnegativity of the solutions. Let **F** = **F**(**x**) denote the vector field representing the right-hand-side of Eqs. (1a)-(1e), with function **x** ≡ (*C, T, B, P, I*). Also, let **n**_*j*_ denote the outward normal unit vector to plane *x*_*j*_ = 0, with *j* = 1, 2, …, 5. That is, **n**_1_ = (−1, 0, 0, 0, 0) and analogously for other **n**_*j*_. Consider the scalar products of the ODE system 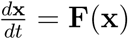 with each **n**_*j*_ and assume that the initial data (*C*(0), *T* (0), *B*(0), *P* (0), *I*(0)) are positive. Then, 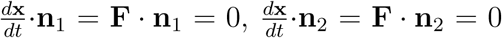 and 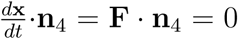 at hyper-surfaces *C* = 0, *T* = 0 and *P* = 0, respectively. Then, the hyper-surfaces *C* = 0, *T* = 0 and *P* = 0 are invariants.

Next, 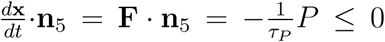 at plane *I* = 0. Finally, 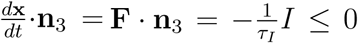 at plane *B* = 0. Hence, pieces of hyper-surfaces 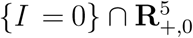 and 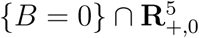 are semipermeable inward 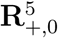.

As a result, 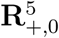 is a positively invariant domain for Eqs. (1a)-(1e). Therefore, nonnegativity of solutions (*C, T, B, P, I*) follows.

Since all parameters entering in Eqs. (1a)-(1e) are finite and the right-hand-side of the system is a continuous function in (*C, T, B, P, I*) in the domain 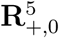, existence of solutions of Eqs. (1a)-(1e) follows from the Cauchy-Peano theorem. Moreover, as the partial derivatives of the right-hand-side of the system are also continuous and bounded in 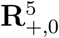, uniqueness follows from the Picard-Lindelöf theorem. This completes the proof.◼

## Appendix B. Basic properties of the reduced mathematical model: system (4)

### Proposition 2.

*For any non negative initial data* (*C*_0_, *T*_0_, *B*_0_) *and all the parameters of the model being positive, the solutions to* *Eqs*. (4) *exist for t >* 0, *are non negative and unique*.

**Proof**. The proof mimics the steps of the proof of Proposition 1 so the repetitive details are omitted.

▪

Before delving into the analysis of system (3), it is convenient to first understand the dynamics of a simplified version of (3) given by Eqs. (4). We begin by calculating the fixed points and determine their stability. These are the points *P*_1_ = (0, 0, 0) and 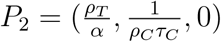.

To analyze the stability of these points, we calculate the Jacobian matrix of Eqs. (4):

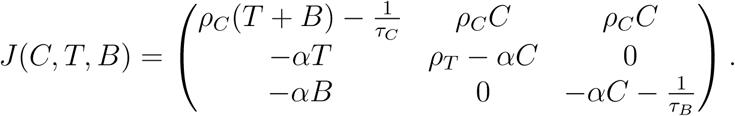

- Equilibrium point *P*_1_ = (0, 0, 0). The Jacobian matrix is

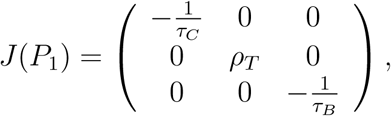

and the eigenvalues are *λ*_1_ = 1*/τ*_*C*_, *λ*_2_ = *ρ*_*T*_ and *λ*_3_ = −1*/τ*_*B*_. Thus, *P*_1_ is a saddle point and therefore, an unstable equilibrium point.
- Equilibrium point 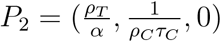. If we make the following linear change of coordinates *x* = *C* − *ρ*_*T*_ */α, y* = *T*−1*/ρ*_*C*_*τ*_*C*_ and *z* = *B* we move the point *P*_2_ to the origin and the system (4) becomes

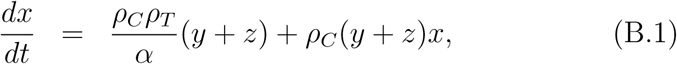

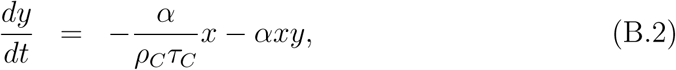

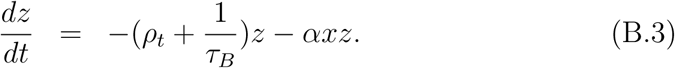 The Jacobian matrix for this point is:

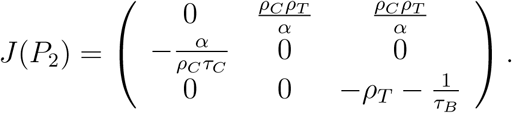

The eigenvalues of the matrix are 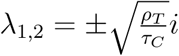 and 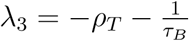.

Since *λ*_1,2_ are imaginary eigenvalues, this point is a non-hyperbolic point. This means that for the linearized system, *P*_2_ is a centre, but it is not possible to conclude its stability for the nonlinear system.

On the other hand, since *λ*_3_ *<* 0, *P*_2_ possesses a local stable manifold corresponding to that eigenvalue. Thus, *P*_2_ has a local center manifold (corresponding to the eigenvalues *λ*_1,2_) of dimension 2 and a local stable manifold (corresponding to the eigenvalue *λ*_3_).

Since *λ*_3_ is negative, all the orbits starting near the equilibrium point approach the center manifold. It is straightforward (although somewhat tedious) to verify that the center manifold is given by *z* = *h*(*x, y*) = 0. So, the qualitative behaviour of the local flow can then be determined from the flow of the following system on the center manifold *z* = 0:

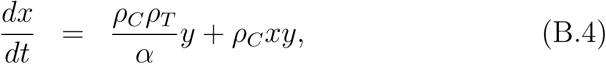

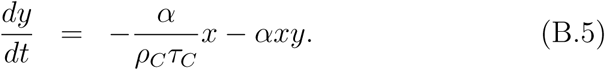

Making the following change of variables

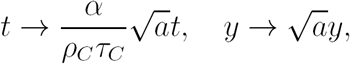

where 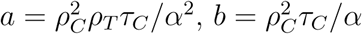 and *m* = *ρ*_*C*_*τ*_*C*_, we obtain

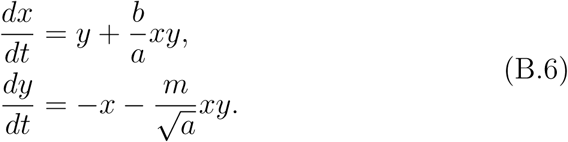

If we introduce polar coordinates, defined by

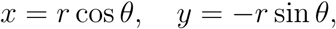

system (B.6) becomes

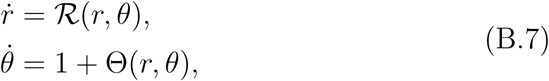

where

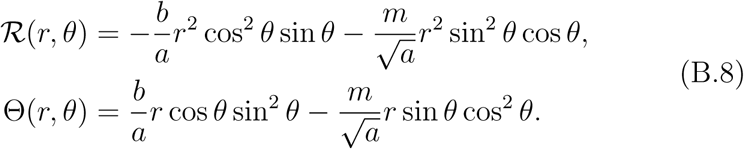

Thus, we can derive an Equation for *r* as a function of *θ* through the differential equation

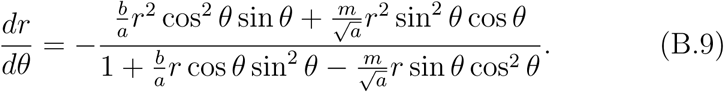

In a neighborhood of *r* = 0,

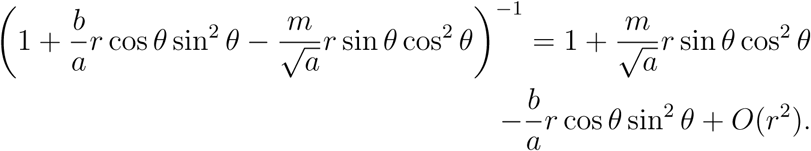

As a result, for small *r* we get the following expression as the Taylor series of Eq. (B.9):

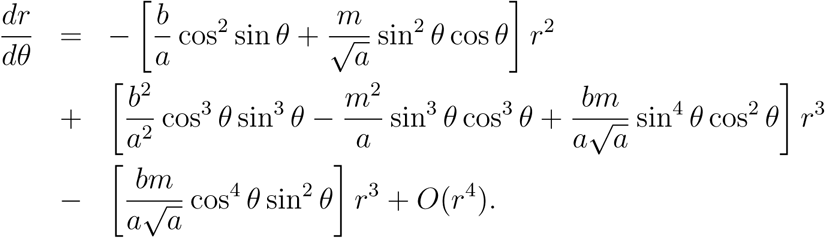

Now, we resort to the method of averaging to perform a change of variable with the effect of reducing the nonautonomous differential Equation into an autonomous one. Let the transformation be

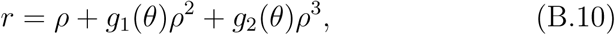

**Figure B.11:**
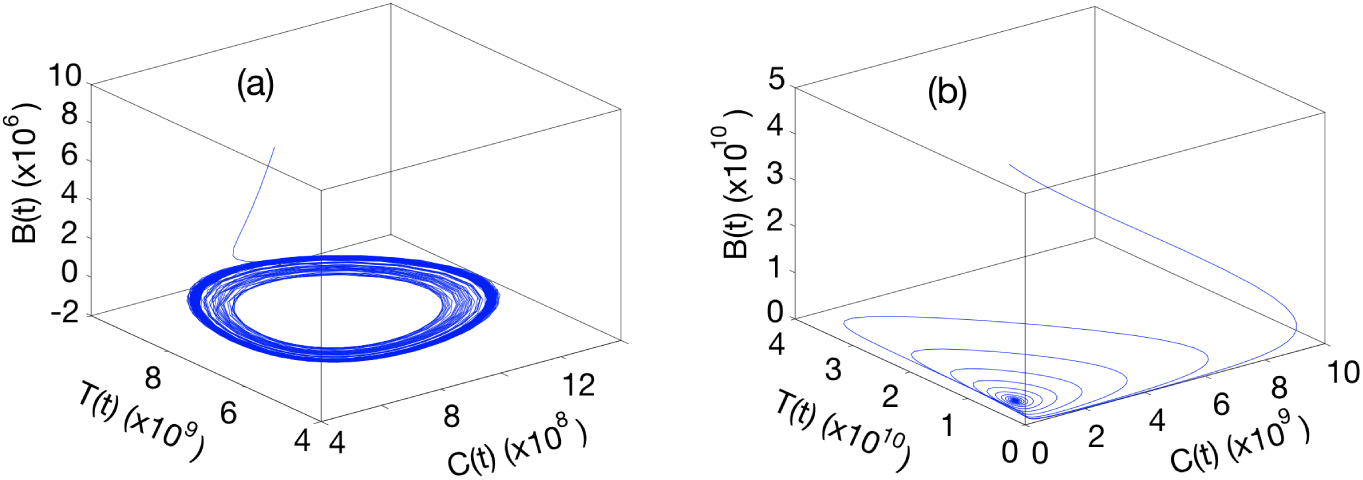
(Left) An orbit of the phase portrait of system (4). (Right) An orbit of the phase portrait of system (3). The parameters used to calculate such orbits are given in Table 1.

with

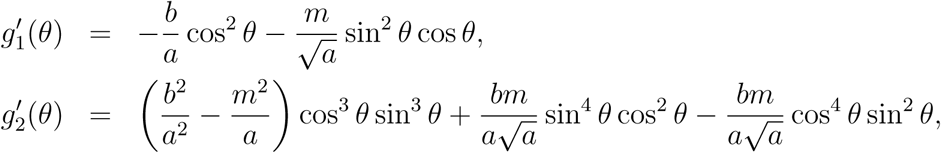

and formally arrive at the equation

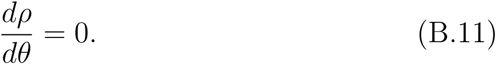

At this point, we are in disposition to apply the Center Theorem of Lyapunov (see for instance [51]) which ensures that when *dr/dθ* can formally be transformed to zero, the equilibrium point is a center. Thus, the origin is a center and thus, the equilibrium point *P*_2_ is also a center.

It is also possible to find a Lyapunov function of the system (B.6) as

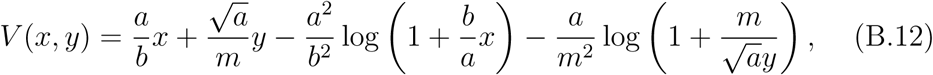

and 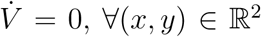. As 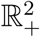 is a positively invariant manifold, all the orbits go periodically around *P*_2_.

An orbit of the phase portrait for a solution of system (4) is shown in Fig. B.11 (Left).

Finally, let (*C*(*t*), *T* (*t*)) be an arbitrary solution of (4) for *B* = 0, and denote its period by 𝒯 *>* 0. It is possible to calculate the time average of variables *C* and *T* (that is, the number of CAR T and tumor cells, respectively). Dividing the first Equation of (4) by *C*, the second by *T* and integrating from 0 to 𝒯 and using the fact that the solutions are periodic, we get

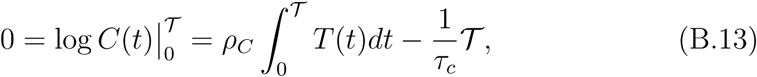

and

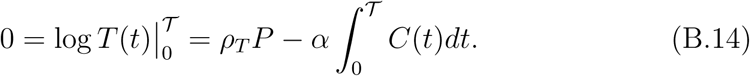

Hence

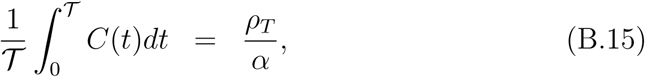

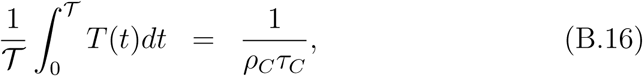

whose values are equal to the equilibrium point *P*_2_ for the two first coordinates.

## Appendix C. Basic properties of the reduced mathematical model: system (3)

We have the following proposition for system (3), which is similar to the previous propositions for systems (1) and (4), and therefore details of the proof are omitted:

### Proposition 3.

*For any nonnegative initial data* (*C*(0), *T* (0), *B*(0)) *and all parameters of the initial value problem given by Eqs. (3a)-(3c) being positive, the solutions for C*(*t*), *T* (*t*) *and B*(*t*) *exist for all t >* 0, *are unique and nonnegative*.

On the other hand, the nonnegative equilibrium points of system (3) are:

- Equilibrium point 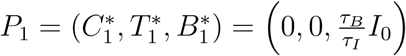.
- Equilibrium point *P*_2_ is given by

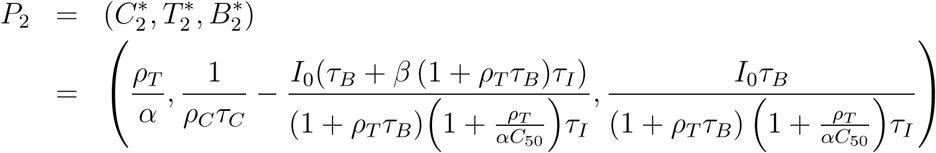

where we assume that

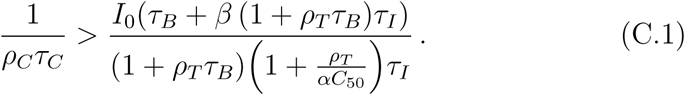
- 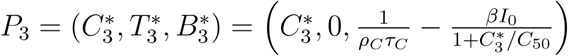 where 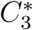 is given by

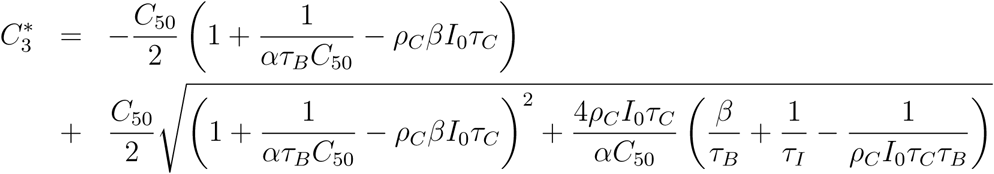

with the following conditions holding

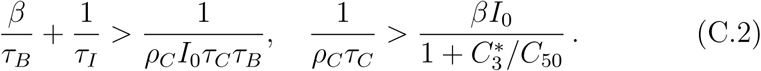
- 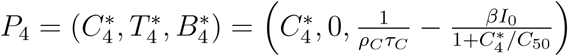 where 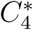 is given by

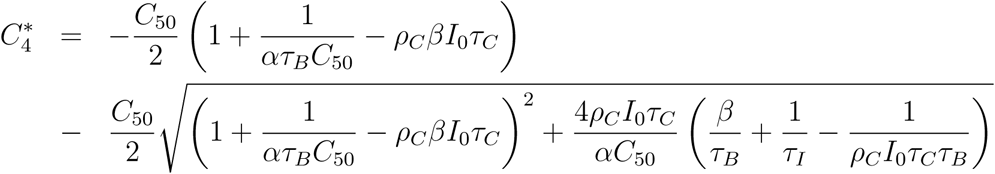

with the following conditions holding

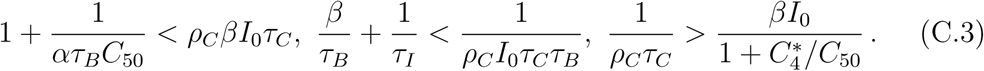

Regarding the study of the stability of the equilibrium points, we obtain the following conclusions:

**Figure C.12:**
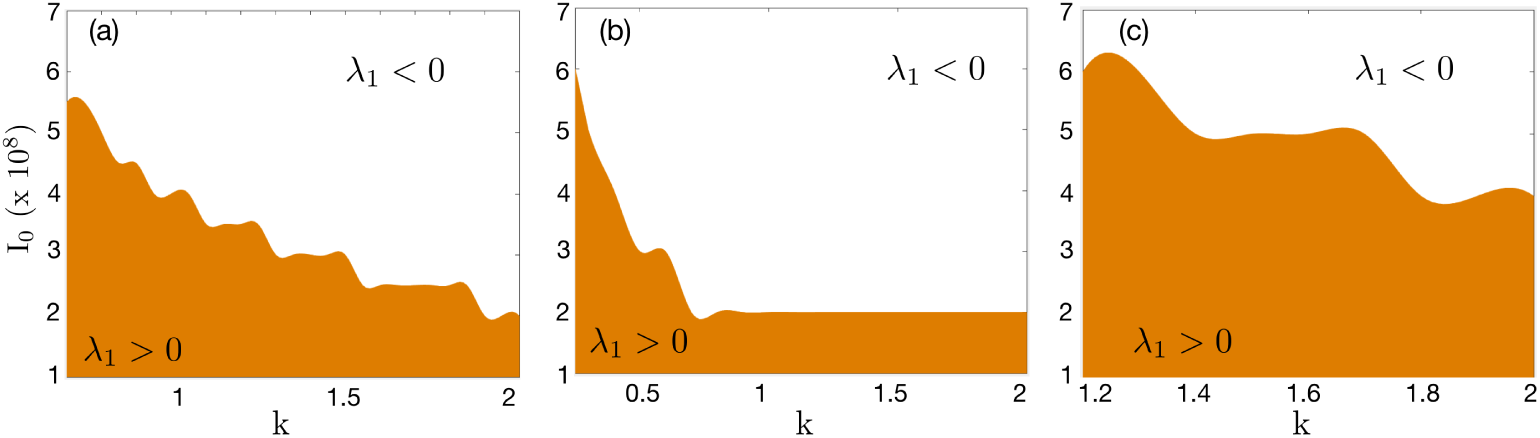
Values of the sign of eigenvalue *λ*_1_ for different values of *I*_0_ and *k* for (a) *α* = 4.5 *·* 10^−11^ (b) *α* = 10^−10^ (c) *α* = 3 *·* 10^−11^. The orange shaded region corresponds to eigenvalue *λ*_1_ positive.

- The eigenvalues of *P*_1_ are

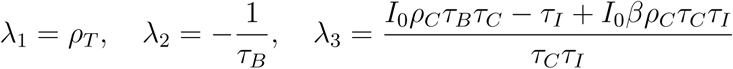

and therefore *P*_1_ is an unstable point (saddle point).
- Using the Routh-Hurwitz criterion, it follows that *P*_2_ is asymptotically stable for any positive *I*_0_ value, assuming that condition (C.1) is satisfied. Since for *I*_0_ = 0 we obtain the system (4), it means that a small perturbation of *I*_0_ = 0, i.e. *I*_0_ = *ε*, with *ε* being sufficiently small, will transform the centers obtained in the system (4) into asymptotically stable foci. Therefore, *I*_0_ = 0 is a bifurcation point since for this value the type of stability changes and we obtain a Hopf bifurcation. An orbit of the phase portrait for a solution of system (3), for this case, is shown in Fig. B.11(right).
- Carrying out a stability study, in a general way, for both *P*_3_ and *P*_4_ is very complex, so we have performed a study within the confines of the parameter range collected in Table 1, which are biologically relevant.

We have observed that for all parameters in Table 1, *P*_4_ has at least one negative component, and thus we do not consider such biologically unfeasible scenarios.

There exist parameters for *P*_3_ for which the components 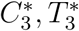 and 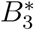 are all positive and correspond to a point that is an asymptotically stable focus. Figure C.12 shows the region where the real eigenvalue, say *λ*_1_, is negative or positive, as a function of *I*_0_ and *k* and for different values of *α*. Figure C.13 depicts all the eigenvalues of *P*_3_ for different values of *I*_0_, *k* and *α*. As it can be seen, the real part of the complex eigenvalues *λ*_2_ and *λ*_3_, is always negative. On the other hand, the real eigenvalue *λ*_1_ changes its sign for different values of *I*_0_, *k* and *α*. Then, the stability and instability of *P*_3_ is given by the sign of *λ*_1_.

**Figure C.13:**
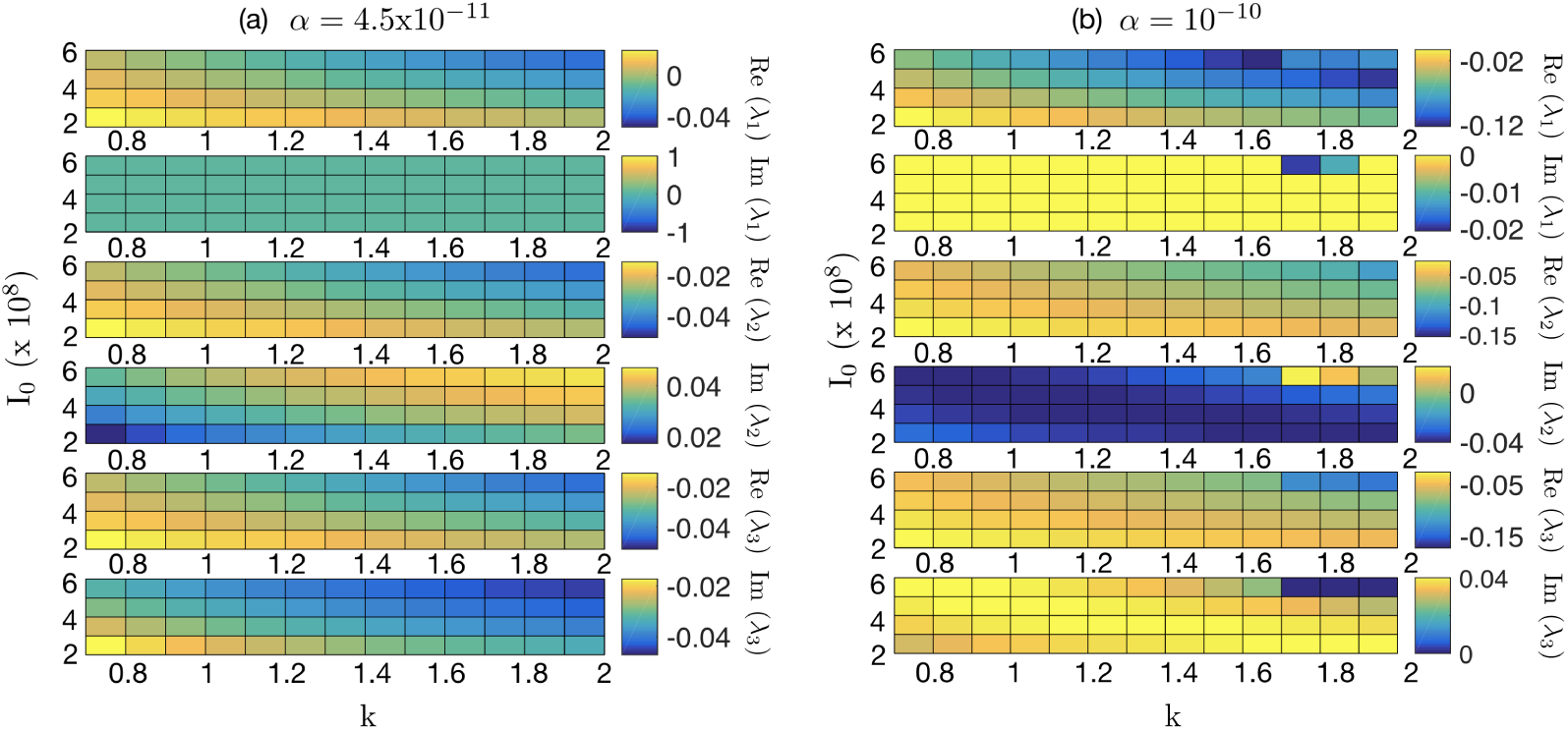
Pseudocolor plots of the real and imaginary parts of *P*_3_ for different values of *I*_0_ and *k* for (a) *α* = 4.5 *·* 10^−11^ and (b) *α* = 10^−10^

## Appendix D. Analytical formulae for system (4)

### Lemma 1.

*Let T*_0_ *and B*_0_ *denote the initial conditions for the tumor and B cells, which are assumed to be positive constants. The exact positive solutions of model Eqs. (4) for the tumor and B cells, denoted by T* = *T* (*t*) *and B* = *B*(*t*), *respectively, satisfy for all t >* 0

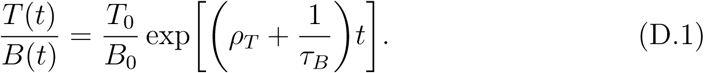

**Proof**. The positive solutions to Eqs. (4b) and (4c) are given, respectively,

by

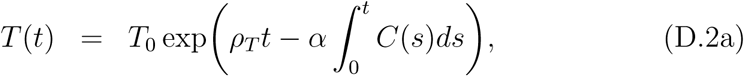

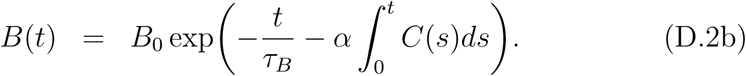

Formula (D.1) easily follows from Eqs. (D.2a) and (D.2b) by calculating their quotient.

### Proposition 4.

*Let t*_*max*_ *denote the time where a local positive maximum of the CAR T cell solution C* = *C*(*t*) *to Eq. (4a) occurs. Then, the positive solutions to Eqs. (4b) and (4c) at t* = *t*_*max*_ *satisfy*

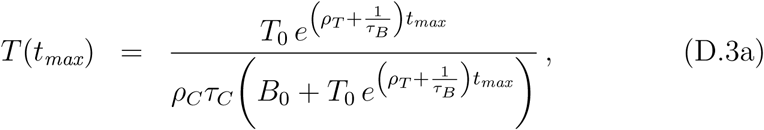

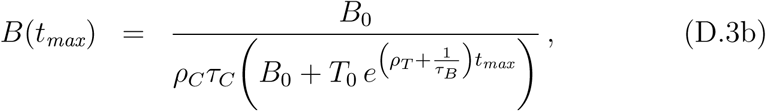

*where T*_0_ *and B*_0_ *are the initial conditions for the tumor and B cells, which are assumed to be positive constants*.

**Proof**. If *C* = *C*(*t*) has a local positive maximum at *t* = *t*_max_, 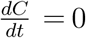 at *t* = *t*_max_. Using Eq. (4a), we get 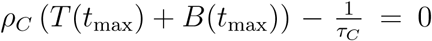. Thus, 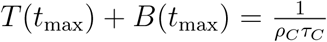. Combining this expression with the above formula (D.1) evaluated at *t* = *t*_max_, Eqs. (D.3) follow.

### Proposition 5.

*Let t*_*max*_ *denote the time where a local positive maximum of the CAR T cell solution C* = *C*(*t*) *to Eq. (4a) occurs. Then t*_*max*_ *can be calculated from the implicit relation*

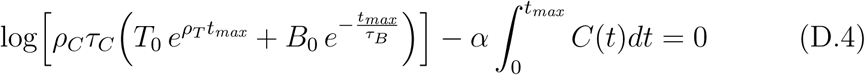

**Proof**. Combining (D.2a) and (D.2b), and setting *t* = *t*_max_, we get

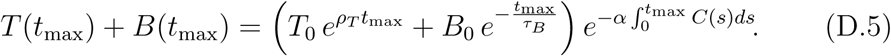

Using the fact that 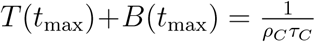, Eq. (D.5) can be finally written as (D.4).

▪

### Proposition 6.

*Let C*_*max*_ *denote the value of the local positive maximum of the CAR T cell solution C* = *C*(*t*) *to Eq. (4a), occurring at time t*_*max*_. *Then*,

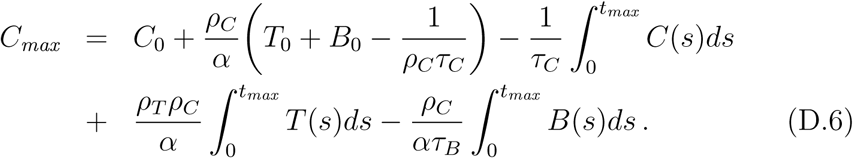

**Proof**. We first combine Eqs. (4) in the form

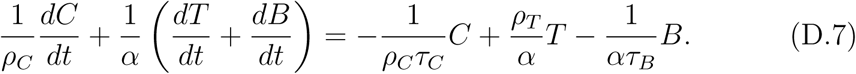

Upon integration, we get

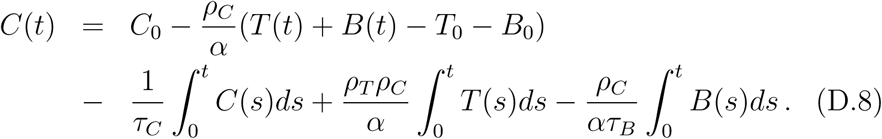

Setting *t* = *t*_max_ and using that 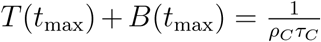 in (D.8), the result follows.

▪

Numerical evaluation of the three integrals in the right-hand-side of (D.8) reveals that, for the parameters collected in Table 1, they are each smaller (by at least one order of magnitude) than the second term (notice that there is also partial cancellation among the three integrals). Hence, we may approximate (D.8) by (6).

